# Assessment of a vancomycin dosing guideline and identification of predictive factors associated with dose and drug trough levels

**DOI:** 10.1101/2022.03.21.22272606

**Authors:** Qingze Gu, Nicola Jones, Philip Drennan, Tim EA Peto, A Sarah Walker, David W Eyre

## Abstract

**Background:** Guidelines and therapeutic drug monitoring are widely used to optimise vancomycin dosing, but their impact remains unclear.

**Objectives:** To assess and optimise vancomycin dosing guidelines in a UK teaching hospital group.

**Methods:** We conducted a retrospective study to evaluate guideline compliance and drug levels following the introduction of a new vancomycin guideline. We used multivariable regression models to investigate factors associated with dosing compliance, drug levels and acute kidney injury (AKI).

**Results:** 3,767 patients received intravenous vancomycin for ≥24h between 01-January-2016 and 01-June-2021. Compliance with recommended loading and initial maintenance doses increased reached 84% and 70% respectively; 72% of subsequent maintenance doses were correctly adjusted. Only 26% first and 32% subsequent levels reached the target range. Drug levels were independently higher in older patients (1.14mg/L per 10 years older with twice-daily dosing [95%CI 1.03,1.25]), with lower eGFR (0.46 mg/L per 10 mL/min/1.73m^2^lower [0.40,0.52]) and higher Elixhauser scores (0.72mg/L per unit higher [0.57,0.88]). For patients with ongoing vancomycin treatment, the conditional probability of achieving target levels at 5 days was >50% and close to 90% at 10 days. Incidence of AKI was low (5.7%), with no evidence that AKI risk increased after guideline implementation (OR=1.09 per year [0.97,1.22], p=0.14). Model estimates were used to propose updated age, weight and eGFR specific guidelines.

**Conclusion:** Despite good compliance with guidelines for vancomycin dosing, the proportion of drug levels achieving the target range remained suboptimal. Adjusting existing guidelines is required to ensure appropriate dosing and better attainment of therapeutic drug levels.

## Introduction

Vancomycin is a glycopeptide antibiotic widely prescribed to treat infections caused by Gram-positive bacteria, including methicillin-resistant *Staphylococcus aureus* (MRSA) and *Enterococcus facium*. Despite advantages such as low cost and established clinical efficacy, vancomycin’s narrow therapeutic index renders appropriate dosing difficult. Vancomycin’s pharmacokinetic/pharmacodynamic (PK/PD) properties mean optimal dosing should maximise the ratio of the 24-hour area under the unbound drug plasma concentration-time curve to minimum inhibitory concentration (AUC/MIC)^1^. An AUC/MIC ratio of 400–600 is recommended to achieve clinical efficacy, while higher levels can cause adverse effects including renal toxicity^2,3^.

Vancomycin trough levels have been widely adopted as a surrogate target for AUC/MIC to simplify clinical management^2^. Individualised dosing and therapeutic drug monitoring (TDM) are necessary to achieve the therapeutic target range, but individual variability and the logistical challenges of coordinating phlebotomy and drug administration in busy hospital settings hinder implementation. Guidelines can facilitate vancomycin dosing and monitoring; many hospitals have developed and implemented local guidelines based on continuously updated national consensus recommendations ^2–5^. However, existing studies demonstrate that guideline implementation has been less effective than expected^6–10^. For example, a recently published study showed that only 37% of vancomycin prescriptions followed recommendations, despite prescribers being aware of guidelines^6^. In another study, despite guidelines and TDM, nearly 70% of patients did not reach therapeutic trough levels during the first three days of treatment^11^. Both incomplete guideline compliance and failure to achieve levels despite following guidance contribute to sub-optimal dosing and may result in antibiotic resistance and increased treatment failure^12^, highlighting the need to further investigate the factors affecting vancomycin dosing and clinical outcomes.

Our hospital group has implemented a new vancomycin dosing guideline since August 2016, increasing the target trough level from 10–15mg/L to 15–20mg/L. The guidelines^13^, delivered via a phone-based app and hospital computers, provide detailed instructions on loading and initial maintenance doses based on the patient’s body weight and renal function, and advise clinicians how to adjust subsequent maintenance doses based on TDM. Implementation is supported by a semi-automatic “powerplan” calculator within the hospital’s electronic patient record system prompting clinicians to prescribe loading and initial maintenance doses based on the guidelines and automatically generating a request for the first vancomycin drug level. This study aimed to investigate the effectiveness of this new vancomycin dosing guideline, identify factors associated with dose and drug levels, and further optimise the guideline accordingly.

## Methods

Data were extracted from the Infections in Oxfordshire Research Database (IORD), containing all admissions to the Oxford University Hospitals (OUH) NHS Foundation Trust in Oxfordshire, United Kingdom. OUH contains 1000 beds in four hospitals, providing secondary care to a population of approximately 600,000 and specialist services to the surrounding region. IORD has approvals from the National Research Ethics Service South Central – Oxford C Research Ethics Committee (19/SC/0403), the Health Research Authority and the national Confidentiality Advisory Group (19/CAG/0144).

Vancomycin is the first-line glycopeptide antibiotic in OUH. The current adult dosing guideline for intravenous vancomycin was implemented on 1 August 2016 (**Table S1**–**S3**). The patient’s actual body weight and estimated glomerular filtration rate (eGFR) determine the loading dose and initial maintenance dose. The first drug trough level should be taken after 48 hours, i.e., before the fourth maintenance dose for twice-daily dosing and before the second maintenance dose for once-daily dosing. Recommendations are included for adjusting subsequent maintenance doses according to trough levels obtained. The target trough level is 15–20mg/L.

We included inpatient treatment courses with intravenous vancomycin lasting ≥24 hours, defining new treatment courses by >14 days between successive doses. Each treatment course contained at least one prescription plus records of individual drug administrations. Patients under 16 years and those admitted to Paediatrics, Paediatric Surgery and Renal Medicine were not covered by the new guideline and so were excluded. We extracted patient characteristics (age, weight, sex, ethnicity, Charlson and Elixhauser scores) and information related to the prescription, administration and monitoring of vancomycin (date and time of prescription and administration, dose, drug trough levels and serum creatinine measurements). Pre-treatment creatinine was the mean over all measurements within two weeks before each treatment course. eGFR was calculated using the Modified Diet in Renal Disease (MDRD) equation^14^.

### Statistical analyses

Regression analyses of different outcomes investigated the new guideline’s effectiveness and examined factors associated with doses and drug levels (details in **Supplementary Methods**; **Table S4**). Compliance of loading doses with the guideline was examined using logistic regression, and resulting first drug trough levels with linear regression. Multinomial logistic and linear regression was used to investigate dose adjustments during maintenance dosing (higher, lower, unchanged) and their impact on subsequent drug levels. The cumulative incidence of reaching the recommended target drug level was investigated using competing risk analysis, and risk factors for AKI were determined using ordinal logistic regression. AKI was defined using the Kidney Disease Improving Global Outcomes (KDIGO) guideline ^15^, with AKI stages 1, 2 and 3 of AKI defined as 1.5– 1.9-fold or ≥26.5μmol/l increase from baseline, 2.0–2.9-fold increase, and ≥3-fold increase or serum creatinine ≥353.6μmol/l, respectively. Regression model findings were used to predict the optimal initial maintenance dose across different patient ages, weights and eGFR.

Continuous explanatory variables were truncated at 1% and 99% to reduce the influence of outliers. Potential non-linear associations were investigated using natural cubic splines. The number of knots was determined based on Akaike Information Criteria (AIC) in univariate models, and then the non-linearity was retained in multivariable models only where it improved model fit (p<0.05). Two-way interactions were included in models where the interaction p<0.05.

## Results

From 1 January 2016 to 1 June 2021, there were 4,573 inpatient vancomycin treatment courses lasting ≥24h in 3,767 patients (**Figure S1**). The median age, weight and eGFR at the start of each course were 62.5 (IQR 48.9–73.2) years, 80.0 (IQR 67.2–93.7) kg, and 90.8 (IQR 70.1–112.2) mL/min/1.73 m^2^, respectively; 58.1% of courses were in males (**Table 1**). Patients had relatively few comorbidities, with most admitted to Trauma and Orthopaedics (57.5%), Neurosurgery (10.5%) and Clinical Haematology (8.7%).

**Table 1.**
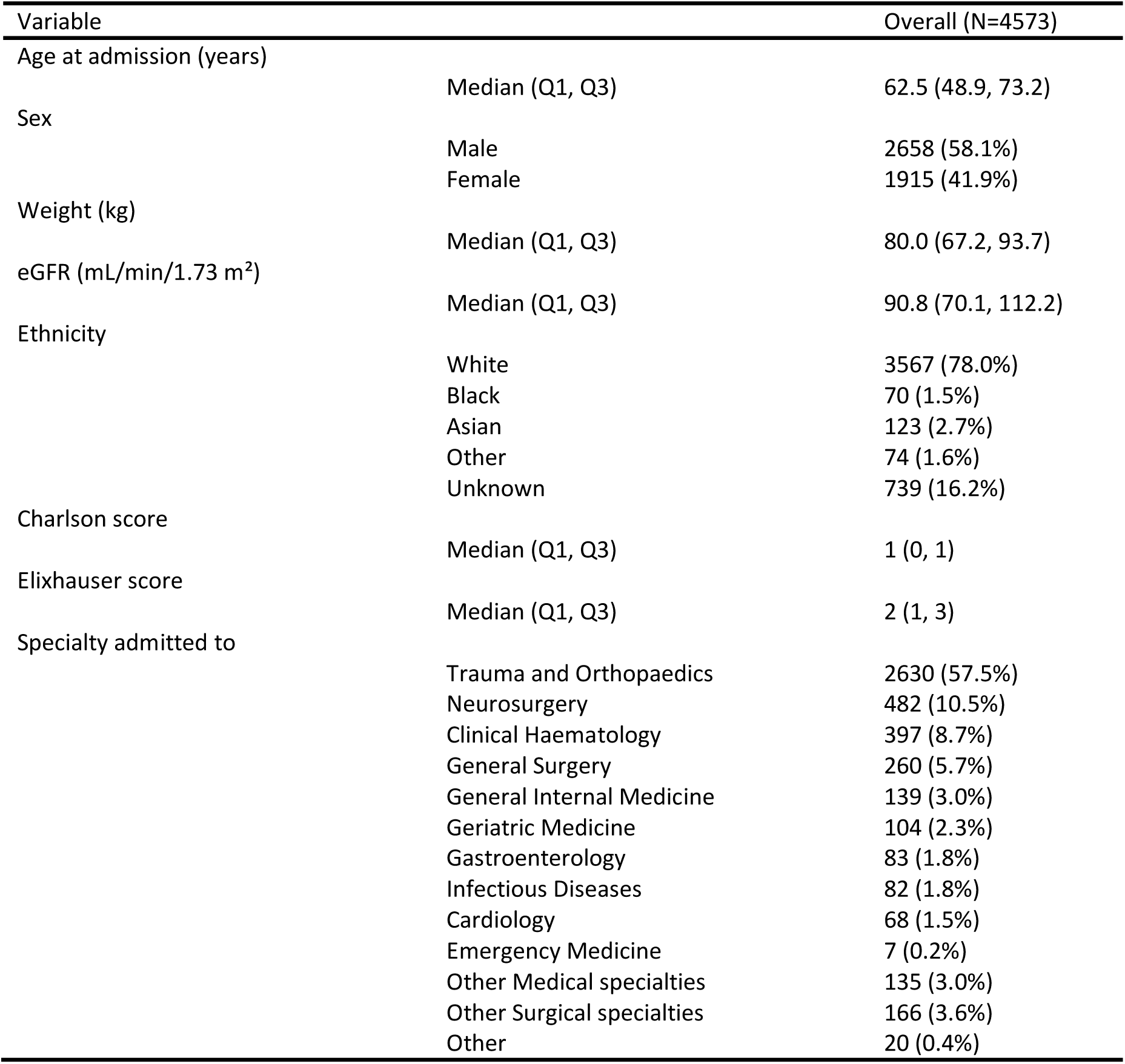
Patient characteristics. Characteristics are shown per treatment course, at the initial prescription for each vancomycin treatment course between 01 January 2016 and 01 June 2021

### Changes in doses following new guideline implementation

Following the implementation of the new vancomycin dosing guideline in August 2016, there were notable shifts in loading doses from predominantly 1000mg (66%) to 2000mg (57%), and in initial maintenance doses which were more varied with the new guideline (**Figure 1A/B/D/E**). Guideline compliance continued to increase slightly over 2017–2021 for both loading and initial maintenance doses (to 84% and 70%, respectively) (**Figure 1G/H**). Although there were multiple independent predictors of loading doses complying with the guideline, the effects were relatively modest (**Table S5, S6, see Supplementary Results**).

**Figure 1.**
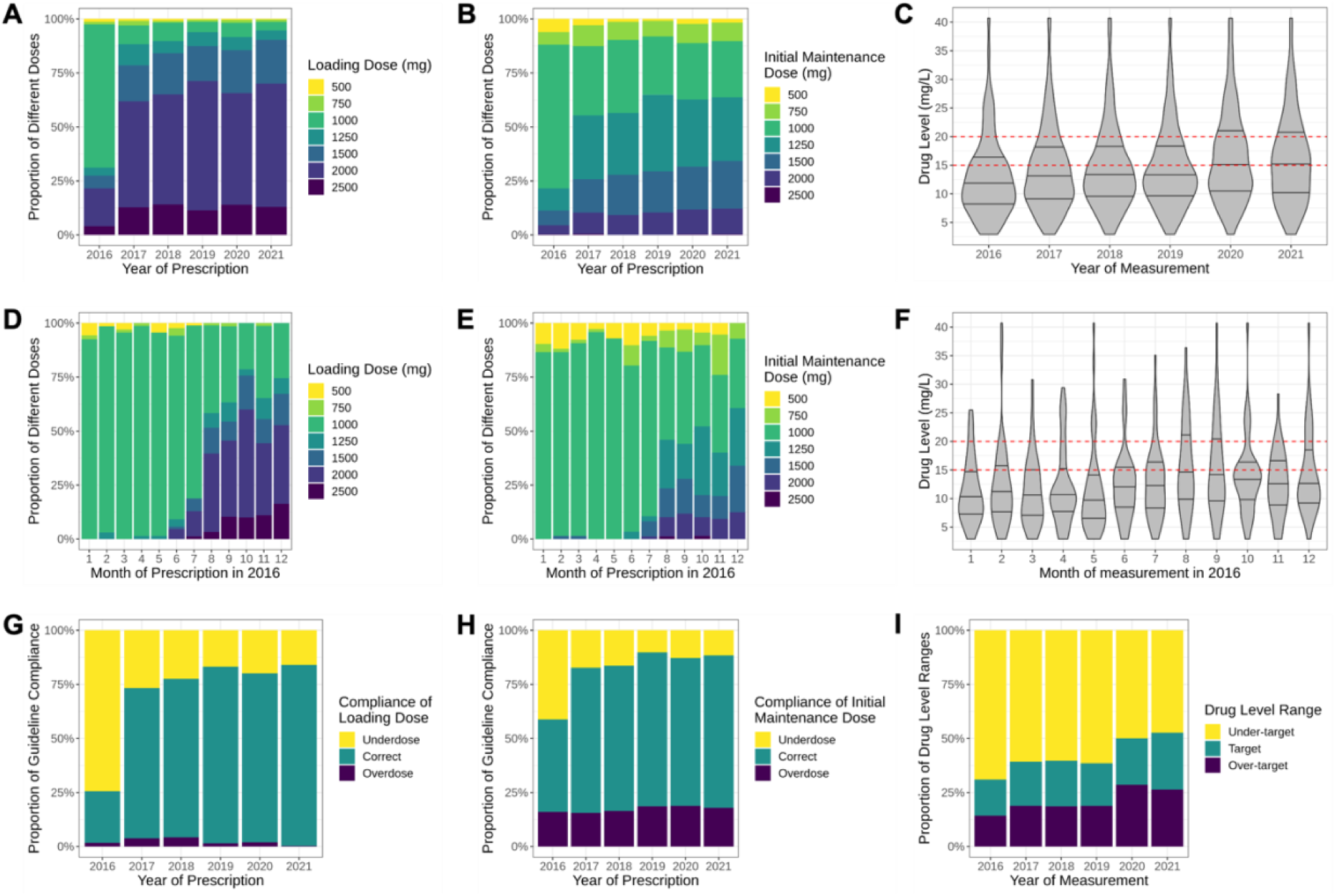
Vancomycin doses and compliance with guidelines. Loading doses, initial maintenance doses and first drug trough levels are shown in panels A–C, by year for 2016 to 2021, and panels D–F by months in 2016. Panels G and H show the proportion of loading and initial maintenance doses compliant with the guidelines by year, and panel I the proportion of drug levels in range (15–20mg/L). The dashed red lines in panel C and F indicate the target vancomycin trough level ranges.

3156 (69%) treatment courses had a drug level taken within 72 hours of starting intravenous vancomycin (median 43.0h (IQR 36.9–47.4) [range 19.6,71.9]). The substantial shifts in loading and initial maintenance dose over time (**Figure 1A/B/D/E**) had relatively small effects on the first drug trough levels (**Figure 1C/F**), with only a modestly increasing trend over 2016–2021 and a limited increase in the proportion of first drug levels reaching the target range (from 17% to 26%, **Figure 1I**). Notably, even in those following the guideline-recommended loading and initial maintenance dose, only 20% of first drug levels reached the target range.

First drug trough levels were independently associated with several baseline factors (**Table S7, S8**) with the strongest effects from eGFR and age rather than doses per kg or dosing compliance. Drug levels were independently lower in those with higher eGFR (0.74mg/L lower for every ten mL/min/1.73m^2^ higher [95% CI 0.62,0.86]). Drug levels were higher in older individuals when initial maintenance doses were administered twice daily (1.12mg/L per 10 years older [0.95,1.28]), with no evidence of the effect of age with once-daily administration (−0.01mg/L [-0.47,0.45, interaction p<0.0001]). Drug levels were also higher in those with higher Elixhauser scores (0.81mg/L per unit higher [0.58,1.03]). As expected for levels obtained ∼48h into treatment, the initial maintenance dose had a stronger effect (0.23mg/L higher per 1mg/kg/day higher [0.16,0.29]) than the effect of the loading dose (−0.06mg/L lower per 1mg/kg/day higher [-0.11,0.01]). Underdosing compared to guideline recommendations in the loading dose and initial maintenance dose resulted in lower drug levels (−0.92mg/L [-1.47,-0.37] and -0.67mg/L [-1.45,0.11], respectively) and conversely overdosing in higher drug levels (1.73mg/L [0.14,3.32] and 1.11mg/L [0.37,1.84], respectively).

### Changes in maintenance doses after initial drug levels

Compared to changes in loading and initial maintenance doses, there were minor changes in the subsequent maintenance doses after guideline implementation (**Figure S3A/B**). Subsequent maintenance doses increased slightly from August 2016 and remained high over 2017–2021. Proportionally, doses within 30–40mg/kg/day rose by about 17%, while doses within 10– 20mg/kg/day fell by about 7% (**Figure S3C**).

For maintenance dose prescriptions issued following measured drug levels (N=4715), 833 (21%) followed a trough level within the target range. Following below target drug levels 2076/2927 (71%) maintenance dose prescriptions increased the dose, and following above target drug levels 706/955 (74%) lowered the dose. Examining the effects of drug levels on subsequent dose adjustments using multinomial logistic regression (**Table S9, S10**), the strongest associations were with most recent eGFR, but these varied according to the previous drug level (**Figure 2**). When the previous drug level was below target (<15mg/L), maintenance doses were generally (72–77% of the time) increased in patients with normal renal function (eGFR≥80mL/min/1.73m^2^), while the likelihood of maintaining or even lowering the current dose increased with lower eGFR. Conversely, high drug levels (>20mg/L) generally (78–80% of the time) led to lower subsequent maintenance doses, although this decreased at higher eGFR (≥80mL/min/1.73m^2^). Effects of other factors were much smaller (**see Supplementary Results**).

**Figure 2.**
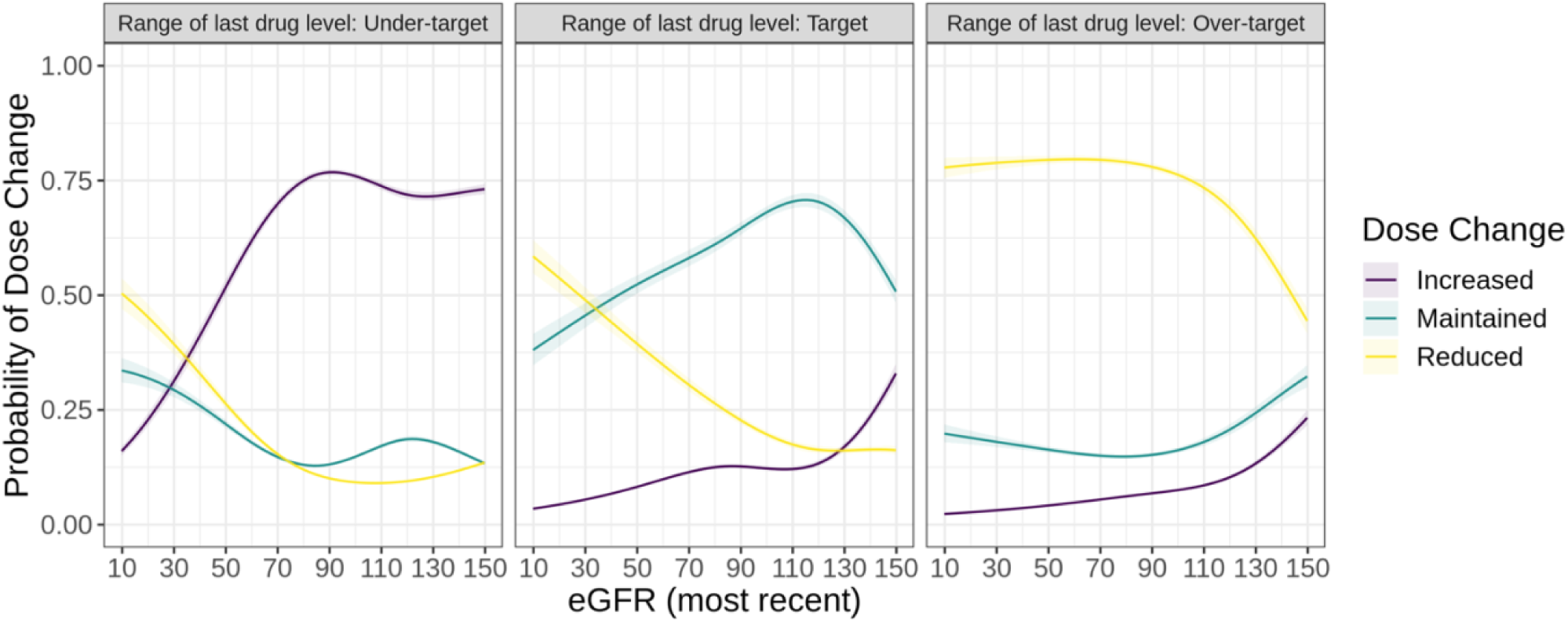
Changes in vancomycin prescriptions following a drug level by drug level (panels) and renal function (x-axis). Under-target, target and over-target are drug levels <15mg/L, 15-20mg/L and >20mg/L respectively. Effects shown are marginalised over the levels of all other factors.

### Subsequent drug levels

Like first drug trough levels, there was also a slightly increasing trend in subsequent drug levels from 2016–2021(**Figure S4**), with an increasing percentage of subsequent drug levels within the target range (from 28% to 32%).

Subsequent drug levels (N=5176) were most strongly associated with maintenance doses and dose adjustments as expected (**Table S11, S12**). Drug levels increased non-linearly with total daily doses (**Figure S5A**), with 20–60mg/kg/day associated with mean levels in range. Adjustments made to maintenance doses in response to drug levels were typically more successful at reducing levels than increasing them: reducing maintenance doses (by a median 7mg/kg/day [IQR 6–12]) typically brought the drug levels within the target range, while increasing doses (by a median 9mg/kg/day [IQR 6–13]) did not (**Figure S5B**). Like initial drug levels, drug levels were higher in older adults when maintenance doses were administered twice daily (1.14mg/L per 10 years older [95%CI 1.03,1.25], with no evidence of an association with age with once-daily dosing (−0.04mg/L [-0.33,0.24, interaction p<0.0001], **Figure S5C**). Drug levels were lower in those with higher eGFR (0.46 mg/L lower per 10 mL/min/1.73m^2^ higher [0.40,0.52]) and were higher in those with higher Elixhauser scores (0.72mg/L per unit higher [0.57,0.88]). As expected, drug levels were lower the longer the time from the last dose to the drug level measurement (**Figure S5D**), with significant variability in the timing of trough levels which did not always follow the recommended timeframe (12h for twice-daily dosing, 24h for once-daily dosing).

### Time to reach therapeutic levels

There was no evidence that higher loading doses per kg led to a higher cumulative incidence of reaching the target level within 72 hours (p=0.47, **Table S13, S14, Figure 3A**), although they did appear to increase the early probability of reaching target levels (within 40 hours). Over the longer term, the probability of reaching the target before stopping vancomycin was higher in the low and medium loading dose groups (p=0.002). Higher loading doses were also associated with a higher cumulative incidence of vancomycin discontinuation (p=0.0001).

**Figure 3.**
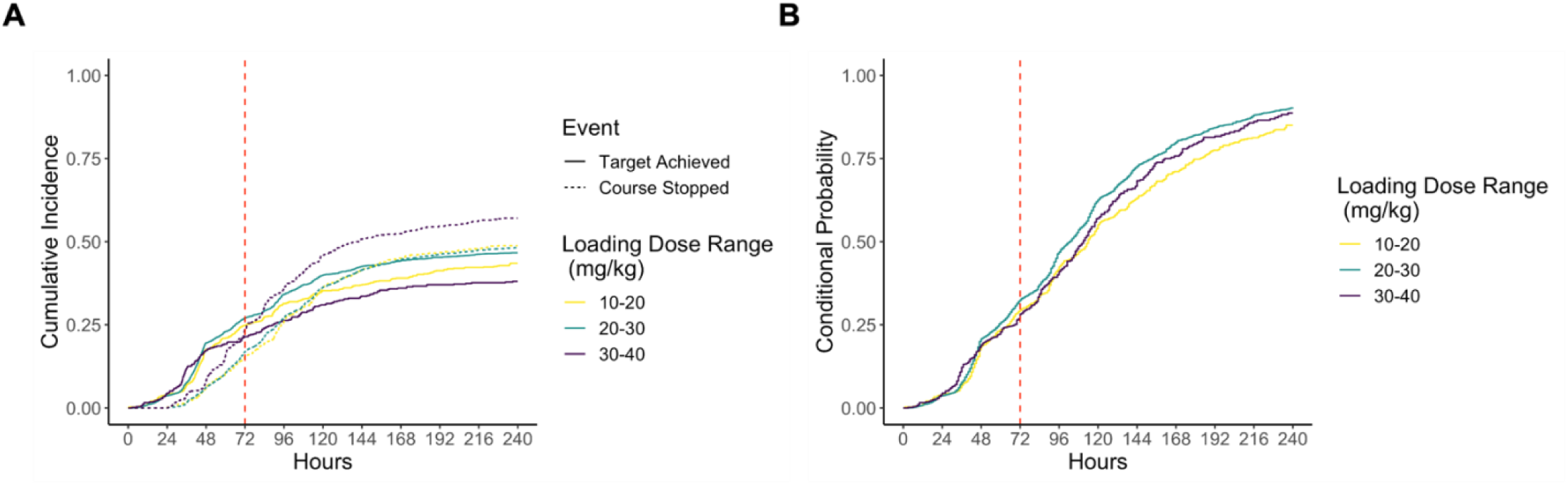
Cumulative incidence of achieving the target trough level. Panel A shows the cumulative incidence of achieving the target trough level (solid line) versus stopping vancomycin before being observed to reach the target (dashed line). Panel B shows the probability of achieving the target conditional on remaining on vancomycin. Both plots are shown according to loading dose 10-20mg/kg (red), 20-30mg/kg (green), 30-40mg/kg (blue). Follow-up time was censored at 240 hours.

In those remaining on vancomycin, the conditional probability of achieving target levels at 5 days was >50% regardless of loading dose; it was similar in the medium and high loading dose groups at 10 days (90%), slightly higher than the low dose group (85%) (**Figure 3B**).

### Acute kidney injury

The risk of nephrotoxicity was relatively low, with only 9 (0.3%), 29 (0.9%) and 147 (4.5%) Stage 3, Stage 2 and Stage 1 AKI cases respectively in 3252 courses with post-treatment creatinine measurements (**Table S15**). Where AKI occurred (n=185), at the end of treatment 50% (93/185) patients had recovered to within ≤1.5 times their pre-treatment creatinine level; for those cases with data recorded within six months (n=101), 88% had recovered (89/101). Higher average drug levels were associated with an increased risk of mild AKI (**Figure 4A/B, Table S16**). AKI risk was also higher in those with lower pre-treatment eGFR and higher Elixhauser scores (**Figure S6, Table S16**). There was no evidence of a change in AKI rate over calendar time after guideline implementation after adjusting for baseline characteristics (odds ratio=1.09 per year, 95%CI=0.97–1.22, p=0.14).

**Figure 4.**
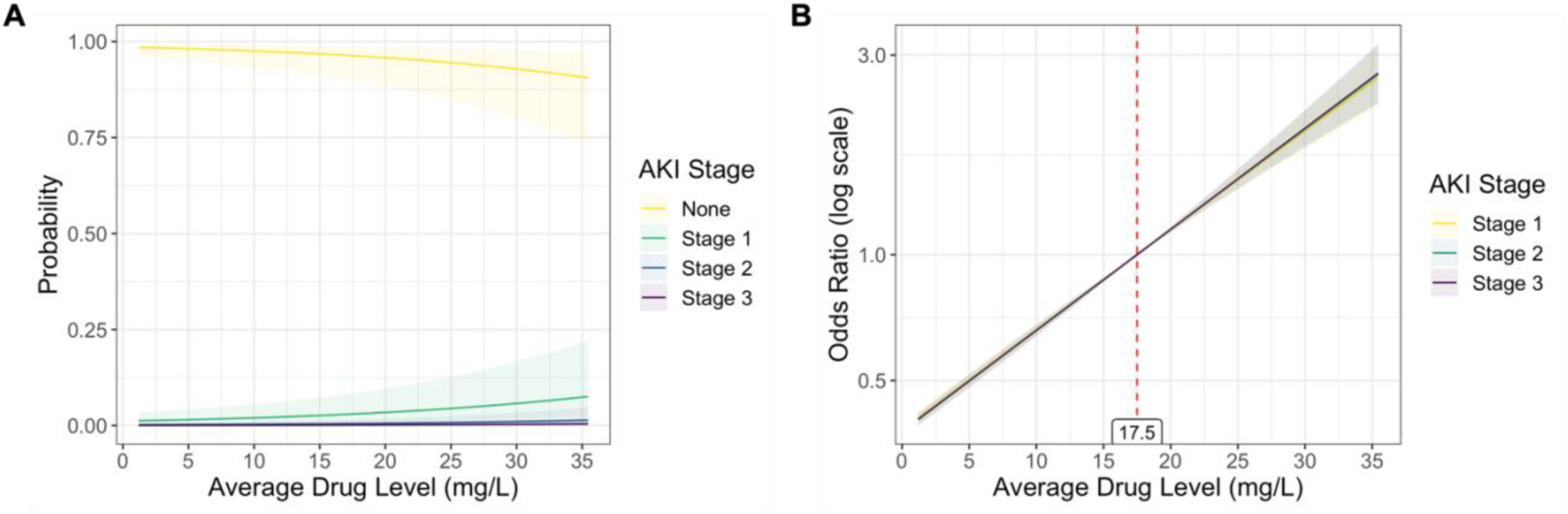
Associations between the probability of different stages of AKI and average drug levels. Panel A shows the probability of stage 1, 2 and 3 AKI, and panel B the odds ratios for AKI (centred at 17.5mg/L). Other predictors were held constant at their mean (for continuous variables) or reference levels (for categorical variables).

### Proposed guideline update

Based on a regression model for the relationship between initial maintenance doses and first drug trough levels in patients of different ages groups, eGFR and body weight, the initial maintenance dose required to achieve the target drug levels in the younger patients (40–60 years) was predicted to be 500–1500 mg higher than the daily dose recommended by the current guideline (**Figure 5**). The predicted optimal dose for patients aged 60–80 years was similar to the current guideline recommendation in patients with eGFR below 90 mL/min/1.73 m^2^, but was higher than the guideline dose in patients with eGFR above 90 mL/min/1.73 m^2^. For patients 60–80 years and ≥110 kg, the predicted optimal dose was lower than the current guideline-recommended dose. New initial maintenance dosing recommendations, based on the current loading dose, are presented in **Table S18**.

**Figure 5.**
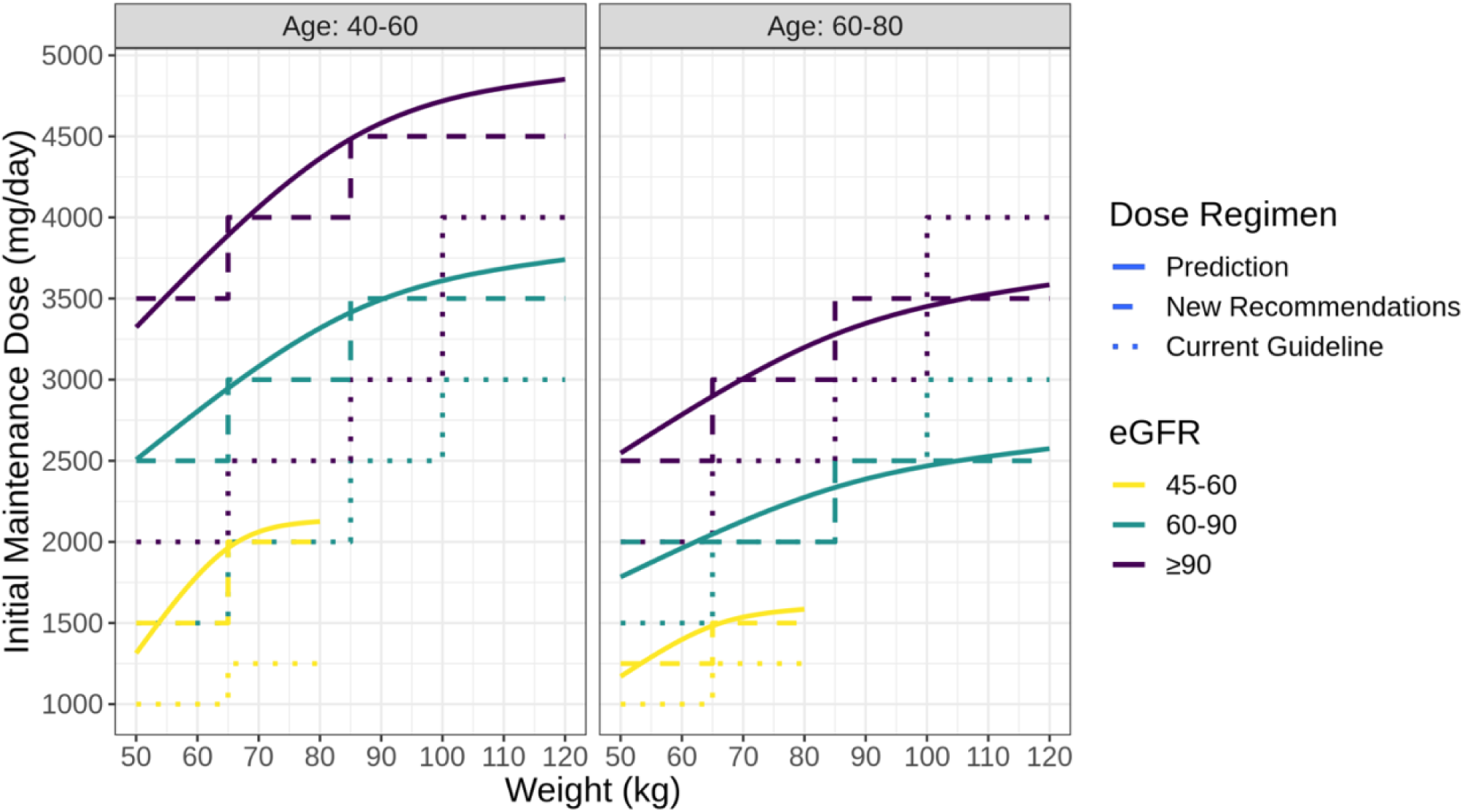
Proposed updated initial maintenance doses by age group, weight and renal function. Model predictions are shown as a solid line, dosing recommendations rounded to doses that can be reliably administered are shown as a dashed line. The current guideline is shown as a dotted line. Dose predictions were not made for patients aged less than 40 years, with eGFR less than 45 mL/min/1.73 m^2^ and in some weight ranges due to a lack of sufficient amounts of data.

## Discussion

Using five years of vancomycin data we show implementing new guidelines increased dosing successfully, but had a more limited impact on achieving therapeutic drug levels. Loading and initial maintenance dose prescriptions showed rapid and good compliance with the new guidelines. Subsequent maintenance doses also adhered well to the guideline recommendations, with 72% of prescriptions correctly adjusting the dose when the drug level was outside the target range. However, the proportion of drug levels reaching the target range was suboptimal (26% initial trough levels and 32% subsequent trough levels), with drug levels being independently higher in patients who were older, had lower eGFR and more comorbidities. For patients with ongoing vancomycin treatment, the conditional probability of achieving the target at 5 days was >50% for all loading dose groups and close to 90% at 10 days. AKI incidence was 5.7%, but for those AKI cases with data recorded within six months, 88% recovered to within 1.5-times their baseline creatinine.

Several previous studies have reported limited vancomycin dosing guideline compliance^6–10^. Supported by a web and phone-based app and electronic patient record prompts, we found that most clinicians in our hospitals followed the new guideline within a few months; compliance with recommended loading dose and first maintenance doses reached 70–80% (**Figure 1D/E/G/H, Table S5**). Around 75% of dose adjustments followed the guidelines, based on the results of drug monitoring and patient characteristics such as initial renal function (**Figure 2**).

Despite relatively good compliance with the guidelines, which were tailored to patients’ weight and eGFR, the proportion of trough levels reaching the target range was low. Only 20% of first drug levels achieved the target even when the guideline was followed, with most patients under-dosed, suggesting that current guidelines may need revision or to account for other patient factors, including age. First and subsequent trough levels were independently higher in older patients with lower eGFR and more comorbidities, and were below target in younger individuals with better renal function and fewer comorbidities. (**Table S7–8, S11–12**).

Achieving higher first trough levels may require increasing initial doses. Theoretically, higher loading doses may help initial infection control and more rapidly achieve minimum vancomycin concentrations (e.g., 10mg/L) needed to prevent the emergence of antibiotic resistance^16^. However, trough drug levels at steady state are more related to initial maintenance doses^17^. We found no evidence that higher loading doses increased the percentage achieving target levels by 72 hours, although there was some evidence of increased levels within 40 hours. Therefore, we propose updating initial maintenance dosing to optimise drug levels and accounting for patient age, which is not considered at present. Using regression model predictions suggests patients 40–60 years receive higher maintenance doses than currently recommended (by 500–1500 mg per day) and higher doses than those 60-80 years. We had insufficient data to produce recommendations for patients <40 or >80 years; careful implementation of recommendations for 40–60 and 60–80 years for these groups could be considered. Further changes may be required in younger patients, e.g., a retrospective study of 151 patients revealed that 40% of patients under 40 years of age eventually required more frequent dosing (every 8h) and took longer to achieve target serum levels^18^.

Higher drug levels in elderly patients further underline the need to adjust vancomycin doses for age. Vancomycin has a longer half-life, a larger volume of distribution and lower clearance in older patients, such that the same dosing regimen may result in higher drug levels^19–22^. Therefore, the creatinine-based eGFR calculations may not accurately reflect the true impact of underlying renal function on clearance in elderly patients^23^. Even in older people with calculated normal eGFR, lower doses may need to be considered to avoid high drug levels. Additionally, the lack of relationship between age and drug levels in those on once-daily doses likely reflects their poorer renal function, driving both a once-daily regime and the primary determinant of drug levels for these patients.

For subsequent dose adjustments, intriguingly, we found that drug levels were generally reduced to within the target range after reducing the maintenance dose, whilst increasing maintenance doses did not raise drug levels to the target within 72 hours. Our current recommendation is to increase dosing by ∼25% for those with sub-therapeutic levels; however, this may be too conservative, particularly with concurrently improving renal function as individuals recover from acute infection or augmented renal clearance of vancomycin^24^.

Despite the modest and inconsistent effectiveness of AUC/MIC in predicting clinical outcomes ^25^, the latest consensus guideline suggests transitioning to AUC-guided dosing due to concerns about the increased risk of nephrotoxicity, particularly if trough levels above those actually needed are targetted^3^. However, despite recommending trough levels of 15-20mg/L, and 20–25% of drug levels being above this target, we found an overall low risk of AKI (5.7%), most of which was mild. This is lower than the range reported for vancomycin-induced nephrotoxicity (10–40%)^26–29^. We found no evidence of increased nephrotoxicity after increasing our target trough level from 10–15mg/L to 15–20mg/L, suggesting concerns about nephrotoxicity from targeting 15–20mg/L trough levels may be smaller than previously reported^27,30,31^. However, the transition to AUC-based monitoring may still be beneficial, allowing for more flexible timing of blood sampling during TDM ^6,32,33^, which may otherwise lead to misinterpretation of trough levels and consequent failure of dosing adjustments. Many studies have reported inappropriately timed sample collection^6,32–35^, and our observations also show this.

Vancomycin’s narrow therapeutic window and inter-and intra-individual variability in pharmacokinetics pose an ongoing challenge for dosing optimisation. Bayesian methods, two-point estimates and continuous infusion are considered more effective to facilitate vancomycin TDM but require significant clinical resource support ^36–40^. Teicoplanin, another glycopeptide antimicrobial, is one alternative to vancomycin where it is licensed. It has comparable efficacy and better tolerability and is therefore simpler to dose^41^. Alternate drugs such as daptomycin, ceftaroline, linezolid and many combination therapies offer more options for vancomycin-resistant infections.

The limitations of this study include that it was performed in a single centre based on electronic health records, so there may have been unmeasured confounding factors and generalisability cannot be assumed. Our guidelines were implemented prior to revised consensus guidelines, with recommendations remaining based on trough levels instead of AUC or AUC/MIC. We acknowledge that the effect of loading doses on steady-state levels may be limited. In models where the loading dose is considered a covariate, it remains informative for the relevant study objectives. This study did not assess the effect of concomitant use of other nephrotoxic drugs on AKI, e.g., piperacillin-tazobactam ^42,43^. Additionally, we did not examine the clinical outcomes of patients given the heterogeneity in the indications for treatment ranging from localised orthopaedic device infection to suspected bacteraemia in profoundly immunosuppressed patients.

In summary, good compliance with vancomycin guidelines was achieved with the assistance of a widely used web and phone app and electronic patient record prompts containing a full suite of antimicrobial guidelines and infection advice. New guidelines successfully achieved higher doses of vancomycin administration, but many patients had sub-therapeutic drug levels. We propose that initial maintenance doses be adjusted for age, as well as weight and renal function. We show how routinely collected electronic data can be used at scale to inform pharmacokinetic studies and dosing recommendations. The risk of AKI in our study was relatively low at 5.7%. The narrow therapeutic window of vancomycin poses an ongoing challenge for dosing optimisation, and the impact of existing guidelines needs to be continuously monitored and adjusted to ensure therapeutic drug levels are achieved.

## Data Availability

The data analysed are not publicly available as they contain personal data but are available from the Infections in Oxfordshire Research Database (https://oxfordbrc.nihr.ac.uk/research-themes-overview/antimicrobial-resistance-and-modernising-microbiology/infections-in-oxfordshire-research-database-iord/), subject to an application and research proposal meeting on the ethical and governance requirements of the Database.

## Acknowledgements

This work was supported by the National Institute for Health Research Health Protection Research Unit (NIHR HPRU) in Healthcare Associated Infections and Antimicrobial Resistance at Oxford University in partnership with the UK Health Security Agency (NIHR200915), and the NIHR Biomedical Research Centre, Oxford. DWE is a Big Data Institute Robertson Fellow. ASW is an NIHR Senior Investigator. The views expressed are those of the authors and not necessarily those of the NHS, the NIHR, the Department of Health or the UK Health Security Agency. The funders had no role in study design, data collection and analysis, decision to publish, or preparation of the manuscript.

## Transparency declarations

DWE declares lecture fees from Gilead outside the submitted work. No other author has a conflict of interest to declare.

