## Supplementary material for "Assessment of a vancomycin dosing guideline and identification of predictive factors associated with dose and drug trough levels"

### Supplementary Methods

From 1 January 2016 to 1 June 2021, there were 7,355 inpatient treatment courses with intravenous vancomycin (**Figure S1**). We firstly excluded courses that were missing any of the baseline characteristics (849). We then excluded courses for patients admitted to Paediatrics, Paediatric Surgery and Renal Medicine and patients aged under 16 years as the guideline was explicitly not applicable to these patients. We then excluded courses with less than 24 hours of intravenous vancomycin, most of which had only a single dose and no drug levels measurements (typically used for surgical prophylaxis or initial empirical treatment). We then considered different outcomes, with specific inclusion criteria as shown in **Figure S1**. Predictor variables and the type of regression for each outcome can be found in **Table S4**.

**Regression analyses**

Descriptive analysis was used to demonstrate the changes in the proportion of loading doses, initial maintenance doses, guideline compliance and first drug trough levels before and after guideline implementation. To investigate the compliance of loading doses with the new guideline, we calculated the recommended dose and dosing interval for loading doses and initial maintenance doses and compared them to the actual doses. Non-compliance was defined as the actual dose failing >10% beyond the recommended dose range. Logistic regression was used to examine factors associated with guideline compliance. For each course, the first drug level was matched with patient characteristics and records of the loading dose and initial maintenance dose. Linear regression was conducted to investigate the association between the baseline characteristics, doses and initial trough levels.

We also used descriptive analysis to demonstrate changes in subsequent maintenance doses and drug levels during the study. Multinomial logistic regression was used to investigate the effects of several baseline characteristics and drug levels on dose adjustments. A further linear regression analysis was performed to examine whether dose adjustments for subsequent maintenance doses contributed to better achieving the target drug level.

Ordered logistic regression was used to investigate the risk of nephrotoxicity at different doses and drug levels and changes in the probability of acute kidney injury (AKI) before and after implementing the guidelines. AKI was defined based on the Kidney Disease Improving Global Outcomes (KDIGO) guideline and calculated from pre-treatment and recent creatinine values. We extracted the records with the maximum AKI stage for each course and calculated average drug levels and doses (averaged over time up to that record) using the trapezoidal rule.

Competing risk analysis was conducted to investigate the effects of different ranges of loading dose on the cumulative incidence of reaching the target level within the censored time. We also examined the probability of achieving the target conditional on remaining on vancomycin for different loading dose groups.

To optimise the recommended maintenance dose, we used a simplified linear model (**Table S17**), incorporating three baseline characteristics (i.e., age, eGFR, weight) and the actual initial maintenance dose, to estimate their relationship with the first drug trough level. With this model, we predicted the first drug trough level resulting from all possible combinations of age, eGFR, weight and daily dose, which simulated the relationship between dose and drug levels in different patient groups. We then filtered out combinations with trough levels on the target range (15–20mg/L) to obtain the potential dose required to achieve target levels in patients of different ages, renal function and body weight.

### Supplementary Results

Guideline compliance independently increased with patient age (odds ratio (OR)=1.14 per 10 years higher, [95%CI 1.09,1.20]) and eGFR (OR=1.05 per 10mL/min/1.73 m² higher [1.03,1.08]) but decreased with higher Elixhauser scores (OR=0.91 [0.84,0.98]). Compliance was independently lower in those admitted to Trauma and Orthopaedics (OR=0.30 [0.18,0.49]) and Cardiology (OR=0.37 [0.17,0.78]) compared to General Internal Medicine. Compliance increased significantly in the months before the implementation of the guidelines (OR=4.41 per month [1.96,9.91]) and continued to increase after implementation but at a much slower rate (OR=1.02 [1.02,1.03]). Compliance rose slightly from the beginning of August (the annual start time for each new cohort of junior doctors) to the end of the following July (OR=1.04 per month [1.02,1.06]). Additionally, compliance was higher for prescriptions written around midday than at midnight (**Figure S2**) and was lower for prescriptions written on Mondays (OR=0.58 vs Wednesday [0.45,0.74]).

In a multinomial logistic regression analysis of subsequent dose adjustments, the odds of reducing maintenance doses were higher in patients with higher pre-treatment eGFR (OR=1.10 per 10mL/min/1.73 m² higher [1.05,1.16]), higher body weight (OR=1.09 per 10kg higher [1.04,1.14]) and in older individuals (OR=1.07 per 10 years higher [1.01,1.13]).

### Supplementary Tables

Recommended loading dose for patients with eGFR ≥ 20mL/min/1.73m^2^.

| Actual Body Weight (kg) | Dose |
| --- | --- |
| 35-49 | 1250mg |
| 50-64 | 1500mg |
| 65-100 | 2000mg |
| >100 | 2500mg |

Recommended loading dose for patients with eGFR < 20mL/min/1.73m^2^.

| Actual Body Weight (kg) | Dose |
| --- | --- |
| <35 | 20mg/kg |
| 35-49 | 1000mg |
| 50-64 | 1250mg |
| >65 | 1500mg |

**Table S1. Recommended loading dose.**

Recommended initial maintenance dose for patients with eGFR ≥ 20mL/min/1.73m^2^.

|  | | eGFR  (mL/min/1.73m^2^) | | | | |
| --- | --- | --- | --- | --- | --- | --- |
|  |  | 20-29 | 30-44 | 45-59 | 60-89 | >90 |
| Actual Body Weight  (kg) | 35-49 | 500mg  Once daily | 500mg  Once daily | 750mg  Once daily | 500mg  Twice daily | 750mg  Twice daily |
|  | 50-64 | 500mg  Once daily | 750mg  Once daily | 1000mg  Once daily | 750mg  Twice daily | 1000mg  Twice daily |
|  | 65-84 | 750mg  Once daily | 1000mg  Once daily | 1250mg  Once daily | 1000mg  Twice daily | 1250mg  Twice daily |
|  | 85-100 | 750mg  Once daily | 1250mg  Once daily | 1500mg  Once daily | 1250mg  Twice daily | 1500mg  Twice daily |
|  | >100 | 1000mg  Once daily | 750mg  Twice daily | 1000mg  Twice daily | 1500mg  Twice daily | 2000mg  Twice daily |

Recommended initial maintenance dose for patients with eGFR < 20mL/min/1.73m^2^.

| Actual Body Weight (kg) | Dose |
| --- | --- |
| <35 | 20mg/kg Once only then re-check level |
| ≥35 | 1000mg Once only then re-check level |

**Table S2. Recommended initial maintenance dose.**

| **Trough level** | **Action required** |
| --- | --- |
| Less than 10mg/L | Increase maintenance dose by two weight bands (see Table S2). |
| 10-14mg/L | Increase maintenance dose by one weight bands (see Table S2). |
| 15-20mg/L | Target trough level. Continue with current dose. |
| 21-25mg/L | Reduce maintenance dose by one weight band (see Table S2). |
| Over 25mg/L | In patients with normal renal function, check the time the sample was taken against time of drug administration. If the sample time does not account for high level then:  1. Omit further doses.  2. Monitor level daily until it reaches 20mg/L or below.  3. Re-start IV vancomycin at two weight bands lower than before (see Table S2). |

**Table S3. Interpretation of vancomycin levels and recommended maintenance dose adjustments.**

| **Variables** | **Loading Dose Compliance (logistic)** | **First Drug Levels (linear)** | **Dose Change Direction (multinomial)** | **Subsequent Drug Levels (linear)** | **Reaching drug level target or stopping drug (competing risks)** | **AKI Stage (ordered logistic)** |
| --- | --- | --- | --- | --- | --- | --- |
| Weight (kg) | Weight | * | Weight | * | * | * |
| Age at admission (years) | Age | Age | Age | Age | Age | Age |
| Sex | Sex | Sex | Sex | Sex | Sex | Sex |
| Charlson Score | Charlson Score | Charlson Score | Charlson Score | Charlson Score | Charlson Score | Charlson Score |
| Elixhauser Score | Elixhauser Score | Elixhauser Score | Elixhauser Score | Elixhauser Score | Elixhauser Score | Elixhauser Score |
| eGFR (pre-treatment , mL/min/1.73 m²) | eGFR (pre-treatment) | eGFR (pre-treatment) | eGFR (pre-treatment) | eGFR (pre-treatment) | eGFR (pre-treatment) | eGFR (pre-treatment) |
| eGFR (most recent, mL/min/1.73 m²) |  |  | eGFR (most recent) |  |  |  |
| Drug Level (mg/L) |  |  | Preceding Drug Level within/above/below Target Range |  |  | Drug Level (time-average) ** |
| Daily Dose (mg/kg/day) |  |  |  | Daily Dose (most Recent) |  | Daily Dose (time-average )** |
| Actual Loading Dose (mg/kg) |  | Actual Loading Dose |  |  | Actual Loading Dose |  |
| Actual Initial Maintenance Dose (mg/kg/day) |  | Actual Initial Maintenance Dose |  |  |  |  |
| Dosing Interval |  | Dosing Interval (Initial Maintenance Dose) |  | Dosing Interval (Most Recent Dose) |  |  |
| Guideline Compliance |  | Guideline Compliance (Both Loading Dose and Initial Maintenance Dose) | Guideline Compliance (Both Loading Dose and Initial Maintenance Dose) |  |  |  |
| Time Since Loading Dose |  | Time Since Loading Dose |  |  |  |  |
| Time Since Preceding Dose |  | Time Since Preceding Dose (Initial Maintenance Dose) |  | Time Since Preceding Dose |  |  |
| Time Since Preceding Drug Level Measurement |  |  |  | Time Since Preceding Drug Level Measurement |  |  |
| Dose Change ( mg/kg/day) |  |  |  | Dose Change |  |  |
| Loading dose Prescription characteristics | Prescription day of year, day of week, minute of day |  |  |  |  |  |
| Specialty admitted to | Specialty |  |  |  |  |  |
| Calendar date and pre/post implementation | Calendar date and pre/post implementation |  |  |  |  |  |

**Table S4. Outcomes considered in regression models together with explanatory variables considered.** Note: Shading indicates explanatory variables that are not applicable (e.g., occur after the outcome, are co-linear with or mediate the effects of other variables, or are not potential confounders for this outcome). *Weight not included as daily dose in mg/kg included instead. **Calculated by averaging over time up to the record with maximum AKI stage for each course using the trapezoidal rule.

| **Variable** | **Incorrect (N=1547)** | **Correct (N=3026)** | **Total (N=4573)** | **p value** |
| --- | --- | --- | --- | --- |
| Age at admission (years) |  |  |  | **0.02** |
| Median (Q1, Q3) | 61.2 (47.8, 73.0) | 63.0 (49.4, 73.3) | 62.5 (48.9, 73.2) |  |
| Sex |  |  |  | 0.08 |
| Male | 927 (59.9%) | 1731 (57.2%) | 2658 (58.1%) |  |
| Female | 620 (40.1%) | 1295 (42.8%) | 1915 (41.9%) |  |
| Weight (kg) |  |  |  | 0.76 |
| Median (Q1, Q3) | 79.7 (66.5, 96.7) | 80.0 (68.0, 92.7) | 80.0 (67.2, 93.7) |  |
| eGFR (mL/min/1.73 m²) |  |  |  | **< 0.0001** |
| Median (Q1, Q3) | 88.5 (68.2, 110.0) | 91.8 (71.6, 113.4) | 90.8 (70.1, 112.2) |  |
| Ethnicity |  |  |  | 0.11 |
| White | 1184 (76.5%) | 2383 (78.8%) | 3567 (78.0%) |  |
| Black | 32 (2.1%) | 38 (1.3%) | 70 (1.5%) |  |
| Asian | 38 (2.5%) | 85 (2.8%) | 123 (2.7%) |  |
| Other | 25 (1.6%) | 49 (1.6%) | 74 (1.6%) |  |
| Unknown | 268 (17.3%) | 471 (15.6%) | 739 (16.2%) |  |
| Charlson score |  |  |  | 0.45 |
| Median (Q1, Q3) | 1 (0, 1) | 1 (0, 1) | 1 (0, 1) |  |
| Elixhauser score |  |  |  | 0.82 |
| Median (Q1, Q3) | 2 (1, 3) | 2 (1, 3) | 2 (1, 3) |  |
| Specialty admitted to |  |  |  | **< 0.0001** |
| General Internal Medicine | 39 (2.5%) | 100 (3.3%) | 139 (3.0%) |  |
| General Surgery | 85 (5.5%) | 175 (5.8%) | 260 (5.7%) |  |
| Trauma and Orthopaedics | 1000 (64.6%) | 1629 (53.8%) | 2629 (57.5%) |  |
| Neurosurgery | 112 (7.2%) | 371 (12.3%) | 483 (10.6%) |  |
| Emergency Medicine | 2 (0.1%) | 5 (0.2%) | 7 (0.2%) |  |
| Gastroenterology | 14 (0.9%) | 69 (2.3%) | 83 (1.8%) |  |
| Clinical Haematology | 99 (6.4%) | 298 (9.8%) | 397 (8.7%) |  |
| Cardiology | 32 (2.1%) | 36 (1.2%) | 68 (1.5%) |  |
| Infectious Diseases | 29 (1.9%) | 53 (1.8%) | 82 (1.8%) |  |
| Geriatric Medicine | 30 (1.9%) | 74 (2.4%) | 104 (2.3%) |  |
| Other Medical specialty | 44 (2.8%) | 91 (3.0%) | 135 (3.0%) |  |
| Other Surgical specialty | 59 (3.8%) | 107 (3.5%) | 166 (3.6%) |  |
| Other | 2 (0.1%) | 18 (0.6%) | 20 (0.4%) |  |

**Table S5. Characteristics of loading dose prescriptions by compliance with the guideline (N=4573).**

| **Predictors** | **OR** | **95% CI** | **p value** |
| --- | --- | --- | --- |
| Age (per 10 years) | 1.14 | 1.09 – 1.20 | **<0.0001** |
| Sex |  |  |  |
| Male | Reference |  |  |
| Female | 1.04 | 0.89 – 1.22 | 0.59 |
| Weight (per 10 kg) | 1.03 | 0.99 – 1.07 | 0.16 |
| eGFR (per 10 mL/min/1.73 m²) | 1.05 | 1.03 – 1.08 | **0.0001** |
| Charlson score | 1.05 | 0.94 – 1.17 | 0.37 |
| Elixhauser score | 0.91 | 0.84 – 0.98 | **0.01** |
| Specialty admitted to |  |  |  |
| General Internal Medicine | Reference |  |  |
| General Surgery | 0.53 | 0.30 – 0.92 | **0.03** |
| Trauma and Orthopaedics | 0.30 | 0.18 – 0.48 | **<0.0001** |
| Neurosurgery | 1.20 | 0.68 – 2.06 | 0.52 |
| Emergency Medicine | 1.03 | 0.15 – 20.18 | 0.98 |
| Gastroenterology | 2.15 | 0.89 – 5.72 | 0.11 |
| Clinical Haematology | 1.10 | 0.61 – 1.95 | 0.74 |
| Cardiology | 0.37 | 0.17 – 0.78 | **0.009** |
| Infectious Diseases | 0.51 | 0.25 – 1.06 | 0.07 |
| Geriatric Medicine | 0.93 | 0.45 – 1.92 | 0.84 |
| Other Medical specialty | 1.02 | 0.51 – 2.07 | 0.95 |
| Other Surgical specialty | 0.50 | 0.27 – 0.91 | **0.02** |
| Other | 8.08 | 1.27 – 123.72 | 0.06 |
| Prescription day of week |  |  |  |
| Wed | Reference |  |  |
| Sun | 0.88 | 0.61 – 1.30 | 0.52 |
| Mon | 0.58 | 0.45 – 0.74 | **<0.0001** |
| Tue | 0.77 | 0.60 – 0.99 | **0.04** |
| Thu | 0.83 | 0.65 – 1.07 | 0.15 |
| Fri | 0.74 | 0.59 – 0.93 | **0.01** |
| Sat | 1.00 | 0.71 – 1.43 | 0.98 |
| Prescription minute of day | See **Figure S2** |  | **0.10** |
| Prescription day of year (from August, per 30 days) | 1.04 | 1.02 – 1.06 | **0.0004** |
| Guideline implementation | 9.18 | 3.93 – 22.09 | **<0.0001** |
| Days before guideline implementation (per 30 days) | 4.41 | 2.23 – 11.41 | **0.0003** |
| Days after guideline implementation (per 30 days) | 1.02 | 1.02 – 1.03 | **<0.0001** |

**Table S6. Independent predictors of compliance of the loading dose with the new guideline.** Note: See Table S5 for the distribution of characteristics. Potential confounding variables (i.e., weight, sex, Charlson score, time of the year, week and day when the prescription was written) were included in the analysis regardless of significance. The days of the year when the prescription was written was counted from 01 August to reflect the annual start time for each new cohort of junior doctors. Natural cubic splines were used with 4 knots at the 10th, 33rd, 67th and 90th percentiles to allow for non-linearity in the effects of time of the day of the prescription on guideline compliance. There was no evidence of interaction between factors in the model (p>0.05).

| **Variable** | **Under-target (N=1871)** | **Target**  **(N=642)** | **Over-target (N=643)** | **Total**  **(N=3156)** | **p value** |
| --- | --- | --- | --- | --- | --- |
| Age at admission (years) |  |  |  |  | **< 0.0001** |
| Median (Q1, Q3) | 57.0 (43.2, 69.2) | 69.0 (60.2, 77.0) | 68.3 (57.6, 76.7) | 62.9 (49.1, 72.8) |  |
| Sex |  |  |  |  | **< 0.0001** |
| Male | 1138 (60.8%) | 346 (53.9%) | 327 (50.9%) | 1811 (57.4%) |  |
| Female | 733 (39.2%) | 296 (46.1%) | 316 (49.1%) | 1345 (42.6%) |  |
| Weight (kg) |  |  |  |  | **< 0.0001** |
| Median (Q1, Q3) | 79.0 (66.9, 92.0) | 80.5 (69.5, 96.0) | 83.5 (70.0, 101.2) | 80.0 (68.0, 94.6) |  |
| eGFR (mL/min/1.73 m²) |  |  |  |  | **< 0.0001** |
| Median (Q1, Q3) | 99.0 (81.5, 120.5) | 84.8 (67.1, 102.9) | 77.2 (62.0, 99.0) | 92.2 (73.2, 113.5) |  |
| Ethnicity |  |  |  |  | **0.002** |
| White | 1428 (76.3%) | 526 (81.9%) | 526 (81.8%) | 2480 (78.6%) |  |
| Black | 29 (1.5%) | 3 (0.5%) | 5 (0.8%) | 37 (1.2%) |  |
| Asian | 57 (3.0%) | 12 (1.9%) | 24 (3.7%) | 93 (2.9%) |  |
| Other | 41 (2.2%) | 6 (0.9%) | 10 (1.6%) | 57 (1.8%) |  |
| Unknown | 316 (16.9%) | 95 (14.8%) | 78 (12.1%) | 489 (15.5%) |  |
| Charlson score |  |  |  |  | **< 0.0001** |
| Median (Q1, Q3) | 1 (0, 1) | 1 (0, 1) | 1 (0, 2) | 1 (0, 1) |  |
| Elixhauser score |  |  |  |  | **< 0.0001** |
| Median (Q1, Q3) | 1 (0, 2) | 2 (1, 3) | 2 (1, 3) | 2 (1, 3) |  |
| Loading dose (mg/kg) |  |  |  |  | 0.09 |
| Median (Q1, Q3) | 24.7 (21.4, 27.6) | 24.0 (20.9, 27.0) | 24.3 (21.6, 27.2) | 24.4 (21.4, 27.4) |  |
| Initial maintenance dose (mg/kg/day) |  |  |  |  | **< 0.0001** |
| Median (Q1, Q3) | 30.6 (25.1, 33.7) | 28.6 (23.9, 32.9) | 29.0 (25.0, 34.1) | 29.9 (25.0, 33.6) |  |
| Maintenance dosing interval (hour) |  |  |  |  | **0.0002** |
| 12 | 1720 (91.9%) | 555 (86.4%) | 576 (89.6%) | 2851 (90.3%) |  |
| 24 | 151 (8.1%) | 87 (13.6%) | 67 (10.4%) | 305 (9.7%) |  |
| Loading dose compliance |  |  |  |  | **0.0002** |
| Underdose | 584 (31.2%) | 170 (26.5%) | 153 (23.8%) | 907 (28.7%) |  |
| Correct | 1250 (66.8%) | 460 (71.7%) | 465 (72.3%) | 2175 (68.9%) |  |
| Overdose | 37 (2.0%) | 12 (1.9%) | 25 (3.9%) | 74 (2.3%) |  |
| Initial maintenance dose compliance |  |  |  |  | **< 0.0001** |
| Underdose | 384 (20.5%) | 109 (17.0%) | 64 (10.0%) | 557 (17.6%) |  |
| Correct | 1280 (68.4%) | 409 (63.7%) | 391 (60.8%) | 2080 (65.9%) |  |
| Overdose | 207 (11.1%) | 124 (19.3%) | 188 (29.2%) | 519 (16.4%) |  |

**Table S7. Characteristics of inpatient vancomycin treatment courses by whether the first drug level reached the target trough level (N=3156).**

| **Predictors** | **Coefficients** | **95% CI** | **p value** |
| --- | --- | --- | --- |
| (Intercept) | 9.36 | 6.91 – 11.82 | **<0.0001** |
| Age (per 10 years) | 1.12 | 0.95 – 1.28 | **<0.0001** |
| Sex |  |  |  |
| Male | Reference |  |  |
| Female | 0.76 | 0.28 – 1.24 | **0.002** |
| eGFR (per 10 mL/min/1.73 m²) | -0.74 | -0.86 – -0.62 | **<0.0001** |
| Charlson score | -0.22 | -0.55 – 0.10 | 0.18 |
| Elixhauser score | 0.81 | 0.58 – 1.03 | **<0.0001** |
| Loading dose (mg/kg) | -0.06 | -0.11 – -0.01 | **0.02** |
| Initial maintenance dose (mg/kg/day) | 0.23 | 0.16 – 0.29 | **<0.0001** |
| Loading dose compliance |  |  |  |
| Correct | Reference |  |  |
| Underdose | -0.92 | -1.47 – -0.37 | **0.001** |
| Overdose | 1.73 | 0.14 – 3.32 | **0.03** |
| Initial maintenance dose compliance |  |  |  |
| Correct | Reference |  |  |
| Underdose | -0.67 | -1.45 – 0.11 | 0.09 |
| Overdose | 1.11 | 0.37 – 1.84 | **0.003** |
| Maintenance dosing interval (hour) |  |  |  |
| 12 | Reference |  |  |
| 24 | 8.20 | 4.76 – 11.65 | **<0.0001** |
| Time since loading doses (hour) | -0.05 | -0.10 – 0.01 | 0.09 |
| Time since initial maintenance doses (hour) | 0.04 | -0.01 – 0.10 | 0.12 |
| Age : Maintenance dosing interval of 24h | -0.11 | -0.16 – -0.06 | **<0.0001** |

**Table S8. Associations with first drug level.** Note: See Table S7 for the distribution of characteristics. Weight was not considered as dose in mg/kg/day was included instead. There was strong evidence of an interaction between age and dosing interval of the initial maintenance dose so this was included (p<0.0001), but no evidence of interaction between other factors in the model (p>0.05). Potential confounding variables (i.e., Charlson score, time since loading doses and time since initial maintenance doses) were included in the analysis regardless of significance.

| Variable | Increased  (N=2277) | Maintained (N=1126) | Reduced (N=1312) | Total  (N=4715) | p value |
| --- | --- | --- | --- | --- | --- |
| Age at admission (years) |  |  |  |  | **< 0.0001** |
| Median (Q1, Q3) | 56.7 (42.5, 69.1) | 63.7 (49.6, 74.4) | 66.0 (53.3, 76.0) | 61.0 (46.7, 72.7) |  |
| Sex |  |  |  |  | **< 0.0001** |
| Male | 1411 (62.0%) | 633 (56.2%) | 717 (54.6%) | 2761 (58.6%) |  |
| Female | 866 (38.0%) | 493 (43.8%) | 595 (45.4%) | 1954 (41.4%) |  |
| Weight (kg) |  |  |  |  | **< 0.0001** |
| Median (Q1, Q3) | 78.0 (65.3, 90.0) | 77.8 (66.0, 91.9) | 80.0 (67.0, 96.2) | 78.7 (66.0, 92.0) |  |
| eGFR (pre-treatment, mL/min/1.73 m²) |  |  |  |  | **< 0.0001** |
| Median (Q1, Q3) | 99.3 (79.1, 124.4) | 86.9 (59.4, 110.1) | 83.7 (62.5, 110.1) | 92.9 (69.0, 117.3) |  |
| eGFR (most recent, mL/min/1.73 m²) |  |  |  |  | **< 0.0001** |
| Median (Q1, Q3) | 109.8 (88.4, 140.1) | 94.8 (67.9, 121.9) | 88.5 (64.9, 116.2) | 101.4 (77.0, 130.2) |  |
| Ethnicity |  |  |  |  | 0.49 |
| White | 1711 (75.1%) | 878 (78.0%) | 1013 (77.2%) | 3602 (76.4%) |  |
| Black | 40 (1.8%) | 15 (1.3%) | 17 (1.3%) | 72 (1.5%) |  |
| Asian | 83 (3.6%) | 29 (2.6%) | 35 (2.7%) | 147 (3.1%) |  |
| Other | 37 (1.6%) | 15 (1.3%) | 18 (1.4%) | 70 (1.5%) |  |
| Unknown | 406 (17.8%) | 189 (16.8%) | 229 (17.5%) | 824 (17.5%) |  |
| Charlson score |  |  |  |  | **< 0.0001** |
| Median (Q1, Q3) | 1 (0, 1) | 1 (0, 2) | 1 (0, 2) | 1 (0, 1) |  |
| Elixhauser score |  |  |  |  | **< 0.0001** |
| Median (Q1, Q3) | 1 (0, 2) | 2 (1, 3) | 2 (1, 3) | 2 (1, 3) |  |
| Range of reported drug level |  |  |  |  | **< 0.0001** |
| Under-target (<15mg/L) | 2076 (91.2%) | 462 (41.0%) | 389 (29.6%) | 2927 (62.1%) |  |
| Target (15–20mg/L) | 123 (5.4%) | 493 (43.8%) | 217 (16.5%) | 833 (17.7%) |  |
| Over-target (>20mg/L) | 78 (3.4%) | 171 (15.2%) | 706 (53.8%) | 955 (20.3%) |  |
| Loading dose compliance |  |  |  |  | 0.13 |
| Underdose | 740 (32.5%) | 322 (28.6%) | 395 (30.1%) | 1457 (30.9%) |  |
| Correct | 1482 (65.1%) | 769 (68.3%) | 885 (67.5%) | 3136 (66.5%) |  |
| Overdose | 55 (2.4%) | 35 (3.1%) | 32 (2.4%) | 122 (2.6%) |  |
| Initial maintenance dose compliance |  |  |  |  | **< 0.0001** |
| Underdose | 555 (24.4%) | 207 (18.4%) | 214 (16.3%) | 976 (20.7%) |  |
| Correct | 1432 (62.9%) | 690 (61.3%) | 796 (60.7%) | 2918 (61.9%) |  |
| Overdose | 290 (12.7%) | 229 (20.3%) | 302 (23.0%) | 821 (17.4%) |  |

**Table S9. Characteristics of prescriptions of maintenance doses by direction of dose change (N=4715).**

|  | **Dose Change: Increased** | | | **Dose Change: Reduced** | | |
| --- | --- | --- | --- | --- | --- | --- |
| **Predictors** | **OR** | **95% CI** | **p value** | **OR** | **95% CI** | **p value** |
| Age (per 10 years) | 1.04 | 0.99 – 1.10 | 0.1483 | 1.07 | 1.01 – 1.13 | **0.03** |
| Sex |  |  |  |  |  |  |
| Male | Reference |  |  | Reference |  |  |
| Female | 0.91 | 0.76 – 1.09 | 0.2896 | 1.11 | 0.92 – 1.33 | 0.28 |
| Weight (per 10 kg) | 0.95 | 0.91 – 1.00 | **0.0376** | 1.09 | 1.04 – 1.14 | **0.0005** |
| eGFR (pre-treatment, per 10 mL/min/1.73 m²) | 0.99 | 0.95 – 1.04 | 0.6931 | 1.10 | 1.05 – 1.16 | **0.0001** |
| eGFR (most recent) | See **Figure 2** |  | **<0.0001** | See **Figure 2** |  | **<0.0001** |
| Charlson score | 0.88 | 0.79 – 0.99 | **0.0392** | 0.95 | 0.84 – 1.07 | 0.38 |
| Elixhauser score | 0.95 | 0.88 – 1.04 | 0.2539 | 1.01 | 0.93 – 1.10 | 0.76 |
| Range of reported drug level |  |  |  |  |  |  |
| Target | Reference |  |  | Reference |  |  |
| Under-target | 14.55 | 9.38 – 22.58 | **<0.001** | 1.55 | 1.11 – 2.15 | **0.01** |
| Over-target | 1.59 | 0.79 – 3.21 | 0.1922 | 6.26 | 4.39 – 8.92 | **<0.0001** |
| Loading dose compliance |  |  |  |  |  |  |
| Correct | Reference |  |  | Reference |  |  |
| Underdose | 1.20 | 0.98 – 1.45 | 0.0733 | 1.10 | 0.90 – 1.35 | 0.37 |
| Overdose | 0.89 | 0.53 – 1.48 | 0.6491 | 0.85 | 0.49 – 1.46 | 0.56 |
| Initial maintenance dose compliance |  |  |  |  |  |  |
| Correct | Reference |  |  | Reference |  |  |
| Underdose | 1.20 | 0.96 – 1.50 | 0.1053 | 0.85 | 0.66 – 1.08 | 0.19 |
| Overdose | 0.82 | 0.64 – 1.04 | 0.1036 | 1.18 | 0.94 – 1.49 | 0.16 |
| eGFR (most recent) : Range of reported drug level | See **Figure 2** |  | **<0.0001** | See **Figure 2** |  | **<0.0001** |

**Table S10. Independent predictors of changes in maintenance doses following drug levels.** Note: See Table S9 for the distribution of characteristics. Natural cubic splines were used with 4 knots at the 10th, 33rd, 67th and 90th percentiles to allow for non-linearity in the effects of the most recent eGFR on dose adjustments. There was strong evidence of an interaction between the range of the last drug level and the most recent eGFR so this was included (p<0.0001), but no evidence of interaction between other factors in the model (p>0.05).

| **Variable** | **Under-target (N=2127)** | **Target (N=1712)** | **Over-target (N=1337)** | **Total (N=5176)** | **p value** |
| --- | --- | --- | --- | --- | --- |
| Age at admission (years) |  |  |  |  | **< 0.0001** |
| Median (Q1, Q3) | 55.3 (41.0, 68.0) | 67.3 (55.3, 76.1) | 68.7 (57.3, 77.9) | 63.4 (49.9, 74.1) |  |
| Sex |  |  |  |  | **< 0.0001** |
| Male | 1338 (62.9%) | 950 (55.5%) | 733 (54.8%) | 3021 (58.4%) |  |
| Female | 789 (37.1%) | 762 (44.5%) | 604 (45.2%) | 2155 (41.6%) |  |
| Weight (kg) |  |  |  |  | **0.0002** |
| Median (Q1, Q3) | 79.0 (67.0, 92.4) | 79.0 (68.2, 92.3) | 81.2 (68.4, 96.6) | 80.0 (68.0, 93.2) |  |
| eGFR (mL/min/1.73 m²) |  |  |  |  | **< 0.0001** |
| Median (Q1, Q3) | 100.5 (80.8, 125.1) | 87.7 (68.8, 108.3) | 75.5 (58.0, 100.0) | 90.8 (68.9, 114.3) |  |
| Ethnicity |  |  |  |  | **< 0.0001** |
| White | 1586 (74.6%) | 1381 (80.7%) | 1042 (77.9%) | 4009 (77.5%) |  |
| Black | 43 (2.0%) | 13 (0.8%) | 12 (0.9%) | 68 (1.3%) |  |
| Asian | 78 (3.7%) | 48 (2.8%) | 39 (2.9%) | 165 (3.2%) |  |
| Other | 40 (1.9%) | 24 (1.4%) | 13 (1.0%) | 77 (1.5%) |  |
| Unknown | 380 (17.9%) | 246 (14.4%) | 231 (17.3%) | 857 (16.6%) |  |
| Charlson score |  |  |  |  | **< 0.0001** |
| Median (Q1, Q3) | 1 (0, 1) | 1 (0, 1) | 1 (0, 2) | 1 (0, 1) |  |
| Elixhauser score |  |  |  |  | **< 0.0001** |
| Median (Q1, Q3) | 1 (0, 2) | 2 (1, 3) | 2 (1, 3) | 2 (1, 3) |  |
| Maintenance dose (mg/kg/day) |  |  |  |  | **< 0.0001** |
| Median (Q1, Q3) | 32.5 (26.4, 39.2) | 30.7 (23.1, 37.9) | 27.6 (20.3, 35.5) | 31.2 (23.7, 37.8) |  |
| Maintenance dosing interval (hour) |  |  |  |  | **< 0.0001** |
| 12 | 1967 (92.5%) | 1516 (88.6%) | 1089 (81.5%) | 4572 (88.3%) |  |
| 24 | 160 (7.5%) | 196 (11.4%) | 248 (18.5%) | 604 (11.7%) |  |
| Dose change amount (mg/kg/day) |  |  |  |  | **< 0.0001** |
| Median (Q1, Q3) | 0.0 (0.0, 6.0) | 0.0 (0.0, 0.0) | 0.0 (0.0, 0.0) | 0.0 (0.0, 0.0) |  |
| Time from last dose to current drug level measurement (hour) |  |  |  |  | **< 0.0001** |
| Median (Q1, Q3) | 11.5 (10.3, 14.2) | 11.0 (9.9, 12.7) | 11.1 (9.7, 18.3) | 11.2 (10.0, 14.0) |  |
| Time from last drug level measurement to current one (hour) |  |  |  |  | **< 0.0001** |
| Median (Q1, Q3) | 33.7 (22.8, 48.2) | 37.9 (23.6, 48.5) | 27.5 (22.2, 47.8) | 34.7 (22.9, 48.2) |  |

**Table S11. Characteristics of maintenance doses by whether the subsequent drug levels reached the target trough level (N=5176).** Drug levels measured ≥6 and ≤48 hours since the last dose.

| **Predictors** | **Coefficients** | **95% CI** | **p value** |
| --- | --- | --- | --- |
| (Intercept) | 12.48 | 11.29 – 13.67 | **<0.0001** |
| Age (per 10 years) | 1.14 | 1.03 – 1.25 | **<0.0001** |
| Sex |  |  |  |
| Male | Reference |  |  |
| Female | 0.26 | -0.07 – 0.58 | 0.12 |
| eGFR (per 10 mL/min/1.73 m²) | -0.46 | -0.52 – -0.40 | **<0.0001** |
| Charlson score | -0.10 | -0.32 – 0.13 | 0.40 |
| Elixhauser score | 0.72 | 0.57 – 0.88 | **<0.0001** |
| Maintenance dose (mg/kg/day) | See **Figure S5A** |  | **<0.0001** |
| Maintenance dosing interval (hour) |  |  |  |
| 12 | Reference |  |  |
| 24 | 10.49 | 8.28 – 12.69 | **<0.0001** |
| Dose change amount (mg/kg/day) | See **Figure S5B** |  | **<0.0001** |
| Time from last dose to current drug level measurement (hour) | See **Figure S5D** |  | **<0.0001** |
| Time from last drug level measurement to current one (hour) | -0.01 | -0.02 – 0.00 | 0.05 |
| Age : Maintenance dosing interval | See **Figure S5C** |  | **<0.0001** |

**Table S12. Independent predictors of subsequent drug levels.** Note: See Table S11 for the distribution of characteristics. Weight was not considered as dose in mg/kg/day was included instead. Natural cubic splines were used with 4 knots at the 10th, 33rd, 67th and 90th percentiles to allow for non-linearity in the effects of maintenance doses and time from the last dose to current drug level measurement on subsequent drug levels. Potential confounding variables (i.e., Charlson score, time from preceding doses and time from preceding drug level measurements) were included in the analysis. There was strong evidence of an interaction between the age and the dosing interval so this was included (p<0.0001), but no evidence of interaction between other factors in the model (p>0.05).

| **Variable** | **Target achieved before 72h whilst on vancomycin**  **(N=1109)** | **Course stopped before 72h and before observing a drug level in the target range**  **(N=796)** | **On vancomycin at 72h without having achieved the target**  **(N=2369)** | **Total**  **(N=4066)** | **p value** |
| --- | --- | --- | --- | --- | --- |
| Age at admission (years) |  |  |  |  | **< 0.0001** |
| Median (Q1, Q3) | 68.5 (58.1, 76.8) | 58.8 (44.9, 71.3) | 59.9 (46.2, 71.5) | 62.7 (49.0, 73.2) |  |
| Sex |  |  |  |  | **0.0004** |
| Male | 560 (53.0%) | 421 (59.4%) | 1385 (60.2%) | 2366 (58.2%) |  |
| Female | 496 (47.0%) | 288 (40.6%) | 916 (39.8%) | 1700 (41.8%) |  |
| Weight (kg) |  |  |  |  | 0.25 |
| Median (Q1, Q3) | 80.0 (68.7, 95.2) | 80.0 (68.0, 94.4) | 80.0 (67.1, 93.5) | 80.0 (67.7, 94.0) |  |
| eGFR  (mL/min/1.73 m²) |  |  |  |  | **< 0.0001** |
| Median (Q1, Q3) | 85.5 (65.0, 104.6) | 90.6 (71.8, 111.8) | 94.3 (73.0, 116.9) | 91.3 (70.9, 112.7) |  |
| Ethnicity |  |  |  |  | **0.008** |
| White | 851 (80.6%) | 564 (79.5%) | 1769 (76.9%) | 3184 (78.3%) |  |
| Black | 6 (0.6%) | 11 (1.6%) | 41 (1.8%) | 58 (1.4%) |  |
| Asian | 24 (2.3%) | 17 (2.4%) | 75 (3.3%) | 116 (2.9%) |  |
| Other | 12 (1.1%) | 19 (2.7%) | 37 (1.6%) | 68 (1.7%) |  |
| Unknown | 163 (15.4%) | 98 (13.8%) | 379 (16.5%) | 640 (15.7%) |  |
| Charlson score |  |  |  |  | **0.0002** |
| Median (Q1, Q3) | 1.0 (0.0, 2.0) | 1.0 (0.0, 1.0) | 1.0 (0.0, 1.0) | 1.0 (0.0, 1.0) |  |
| Elixhauser score |  |  |  |  | **< 0.0001** |
| Median (Q1, Q3) | 2.0 (1.0, 3.0) | 1.0 (0.0, 2.0) | 1.0 (1.0, 2.0) | 2.0 (1.0, 3.0) |  |
| Loading dose (mg/kg) |  |  |  |  | **0.006** |
| Median (Q1, Q3) | 24.1 (21.2, 27.3) | 24.8 (21.5, 28.6) | 24.4 (21.0, 27.4) | 24.4 (21.2, 27.6) |  |

**Table S13. Characteristics of vancomycin treatment courses by the trough level outcome.**

| **Predictors** | **HR*** | **95% CI** | **p value** |
| --- | --- | --- | --- |
| Age (per 10 years) | 1.26 | 1.21 – 1.31 | **<0.0001** |
| Sex |  |  |  |
| Male | Reference |  |  |
| Female | 1.18 | 1.04 – 1.33 | **0.01** |
| eGFR (per 10 mL/min/1.73 m²) | 0.98 | 0.96 – 1.00 | **0.04** |
| Charlson score | 1.04 | 0.96 – 1.13 | 0.36 |
| Elixhauser score | 1.00 | 0.94 – 1.06 | **0.91** |
| Loading dose (mg/kg) | 1.00 | 0.99 – 1.01 | 0.47 |

**Table S14. Independent predictors of reaching target vs stopping vancomycin before 72h and before observing a drug level in the target range.** Note: *HR = Hazard Ratio. See Table S13 for the distribution of characteristics. Weight not included as the loading dose in mg/kg included instead. There was no evidence of interaction between factors in the model (p>0.05).

| **Variable** | **None (N=3067)** | **Stage 1 (N=147)** | **Stage 2 (N=29)** | **Stage 3 (N=9)** | **Total (N=3252)** | **p value** |
| --- | --- | --- | --- | --- | --- | --- |
| Age at admission (years) |  |  |  |  |  | 0.07 |
| Median (Q1, Q3) | 62.5 (49.0, 73.1) | 65.1 (51.0, 74.8) | 60.9 (53.5, 68.2) | 54.4 (34.9, 58.5) | 62.6 (49.1, 73.2) |  |
| Sex |  |  |  |  |  | 0.04 |
| Male | 1789 (58.3%) | 69 (46.9%) | 19 (65.5%) | 5 (55.6%) | 1882 (57.9%) |  |
| Female | 1278 (41.7%) | 78 (53.1%) | 10 (34.5%) | 4 (44.4%) | 1370 (42.1%) |  |
| Weight (kg) |  |  |  |  |  | 0.44 |
| Median (Q1, Q3) | 79.7 (67.0, 93.5) | 77.5 (66.2, 90.6) | 77.6 (66.0, 87.7) | 95.0 (80.0, 101.0) | 79.4 (67.0, 93.4) |  |
| eGFR (mL/min/1.73 m²) |  |  |  |  |  | **< 0.0001** |
| Median (Q1, Q3) | 92.1 (71.4, 114.2) | 67.4 (31.1, 110.3) | 85.5 (75.9, 115.8) | 102.1 (85.7, 105.7) | 91.6 (70.1, 113.8) |  |
| Ethnicity |  |  |  |  |  | 0.98 |
| White | 2373 (77.4%) | 114 (77.6%) | 21 (72.4%) | 8 (88.9%) | 2516 (77.4%) |  |
| Black | 47 (1.5%) | 4 (2.7%) | 0 (0.0%) | 0 (0.0%) | 51 (1.6%) |  |
| Asian | 93 (3.0%) | 4 (2.7%) | 1 (3.4%) | 0 (0.0%) | 98 (3.0%) |  |
| Other | 46 (1.5%) | 2 (1.4%) | 0 (0.0%) | 0 (0.0%) | 48 (1.5%) |  |
| Unknown | 508 (16.6%) | 23 (15.6%) | 7 (24.1%) | 1 (11.1%) | 539 (16.6%) |  |
| Charlson score |  |  |  |  |  | **< 0.0001** |
| Median (Q1, Q3) | 1 (0, 1) | 1 (1, 2) | 1 (1, 2) | 1 (0, 1) | 1 (0, 1) |  |
| Elixhauser score |  |  |  |  |  | **< 0.0001** |
| Median (Q1, Q3) | 2 (1, 3) | 2 (1, 4) | 2 (1, 3) | 2 (1, 2) | 2 (1, 3) |  |
| Average dose (mg/kg)* |  |  |  |  |  | 0.36 |
| Median (Q1, Q3) | 16.7 (14.3, 18.9) | 16.7 (14.2, 18.9) | 15.6 (12.2, 18.4) | 18.4 (15.9, 21.9) | 16.7 (14.3, 18.9) |  |
| Average drug level (mg/L)* |  |  |  |  |  | **< 0.0001** |
| Median (Q1, Q3) | 10.8 (7.3, 15.1) | 12.3 (9.2, 17.6) | 13.6 (11.4, 18.5) | 15.3 (10.9, 35.4) | 10.9 (7.4, 15.2) |  |

**Table S15. Characteristics of vancomycin treatment courses by worst acute kidney injury stages (N=3252).** *Average drug levels and doses calculated as a time-average over all values up to and including the creatinine measurement defining the worst acute kidney injury stage, using the trapezoidal rule.

| **Predictors** | **OR** | **95% CI** | **p value** |
| --- | --- | --- | --- |
| Age (per 10 years) | 0.92 | 0.83 – 1.01 | **0.08** |
| Sex |  |  |  |
| Male | Reference |  |  |
| Female | 1.28 | 0.94 – 1.75 | 0.11 |
| eGFR (per 10 mL/min/1.73 m²) | See **Figure S6** |  | **<0.0001** |
| Charlson score | 1.06 | 0.87 – 1.29 | 0.55 |
| Elixhauser score | 1.17 | 1.01 – 1.34 | **0.03** |
| Average dose (mg/kg) | 1.04 | 1.00 – 1.09 | **0.04** |
| Average drug level (mg/L) | 1.06 | 1.03 – 1.08 | **<0.0001** |

**Table S16. Independent predictors of the maximum acute kidney injury stage per vancomycin course.** Note: See Table S15 for the distribution of characteristics. Weight not included as average doses in mg/kg included instead. Natural cubic splines were used with 3 knots at the 10th, 50th and 90th percentiles to allow for non-linearity in the effects of eGFR on the probability of different stages of AKI. No evidence of interaction between factors in the model (p>0.05).

| **Predictors** | **Coefficients** | **95% CI** | **p value** |
| --- | --- | --- | --- |
| (Intercept) | 16.70 | 12.39 – 21.00 | **0.001** |
| Age (per 10 years) | -0.59 | -1.15 – -0.02 | **0.04** |
| eGFR (mL/min/1.73 m²) | Non-linear term |  | **<0.0001** |
| Weight (kg) | Non-linear term |  | 0.85 |
| Initial maintenance dose (per 100 mg/day) | 0.09 | -0.09 – 0.28 | 0.32 |
| Age : Initial maintenance dose | 0 | 0– 0 | **0.0005** |
| Age : eGFR | Non-linear term |  | 0.20 |
| eGFR : Weight | Non-linear term |  | **0.007** |
| Age : Weight | Non-linear term |  | 0.16 |

**Table S17. Associations between first drug trough levels and age, eGFR, weight and initial maintenance doses.** Note: Natural cubic splines were used with two knots at 80 and 105 to allow for non-linearity in the effects of eGFR, two knots at 72 and 89 for weight. Two-way interactions between age, eGFR, weight and initial maintenance doses were included.

| Age: 40-60 years | | eGFR  (mL/min/1.73m^2^) | | |
| --- | --- | --- | --- | --- |
|  |  | 45-60 | 60-90 | >90 |
| Actual Body Weight  (kg) | 50-65 | 750mg  Twice daily | 1250mg  Twice daily | 1750mg  Twice daily |
|  | 65-85 | 1000mg  Twice daily | 1500mg  Twice daily | 2000mg  Twice daily |
|  | 85-100 | * | 1750mg  Twice daily | 2250mg  Twice daily |
|  | >100 | * | 1750mg  Twice daily | 2250mg  Twice daily |

| Age: 60-80 years | | eGFR  (mL/min/1.73m^2^) | | |
| --- | --- | --- | --- | --- |
|  |  | 45-60 | 60-90 | >90 |
| Actual Body Weight  (kg) | 50-65 | 1250mg  Once daily | 1000mg  Twice daily | 1250mg  Twice daily |
|  | 65-85 | 750mg  Twice daily | 1000mg  Twice daily | 1500mg  Twice daily |
|  | 85-100 | * | 1250mg  Twice daily | 1750mg  Twice daily |
|  | >100 | * | 1250mg  Twice daily | 1750mg  Twice daily |

**Table S18. Optimised dosing recommendations for initial maintenance dose for patients aged 40–60 years and 60–80 years.** *Dose predictions were not made for patients in some weight and eGFR ranges due to a lack of sufficient amounts of actual data.

### Supplementary Figures

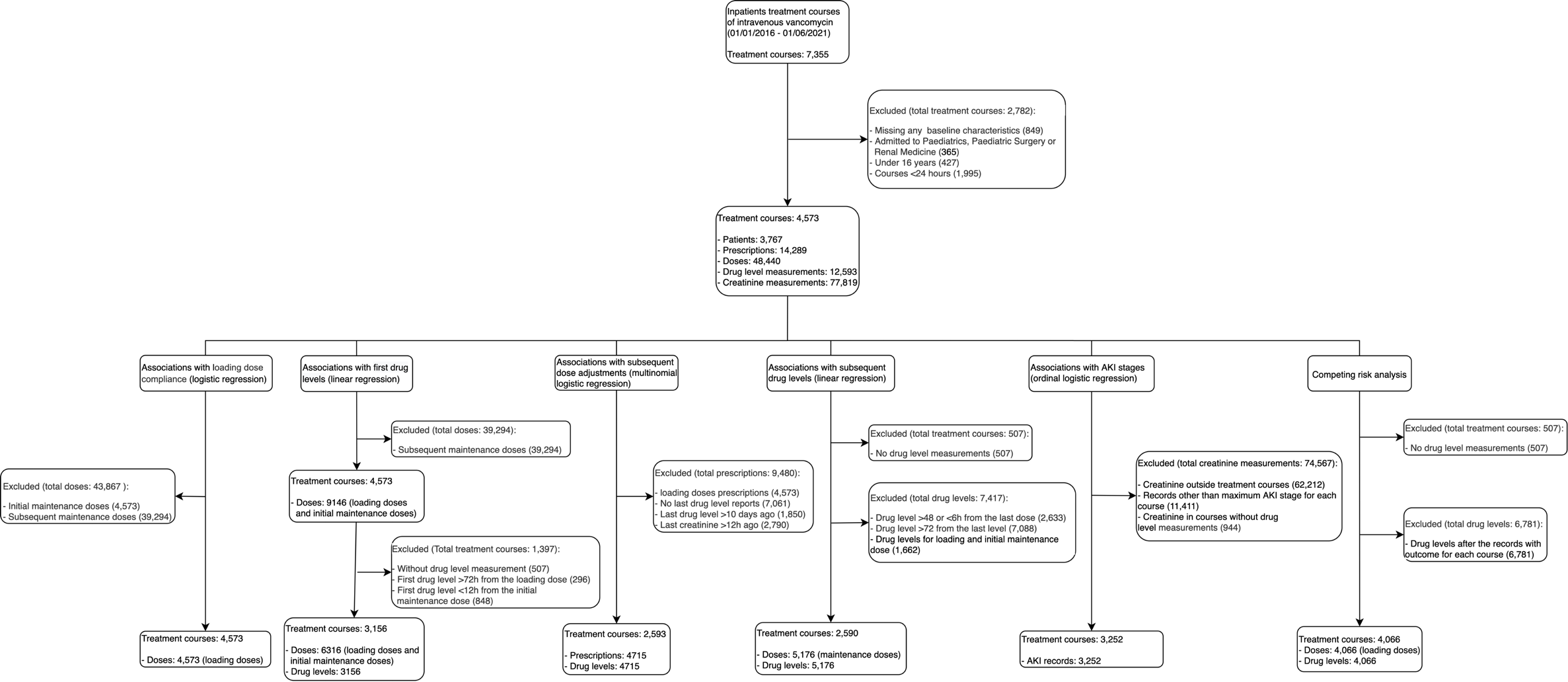

**Figure S1. Flowchart of the dataset selection and generation for each outcome investigated**

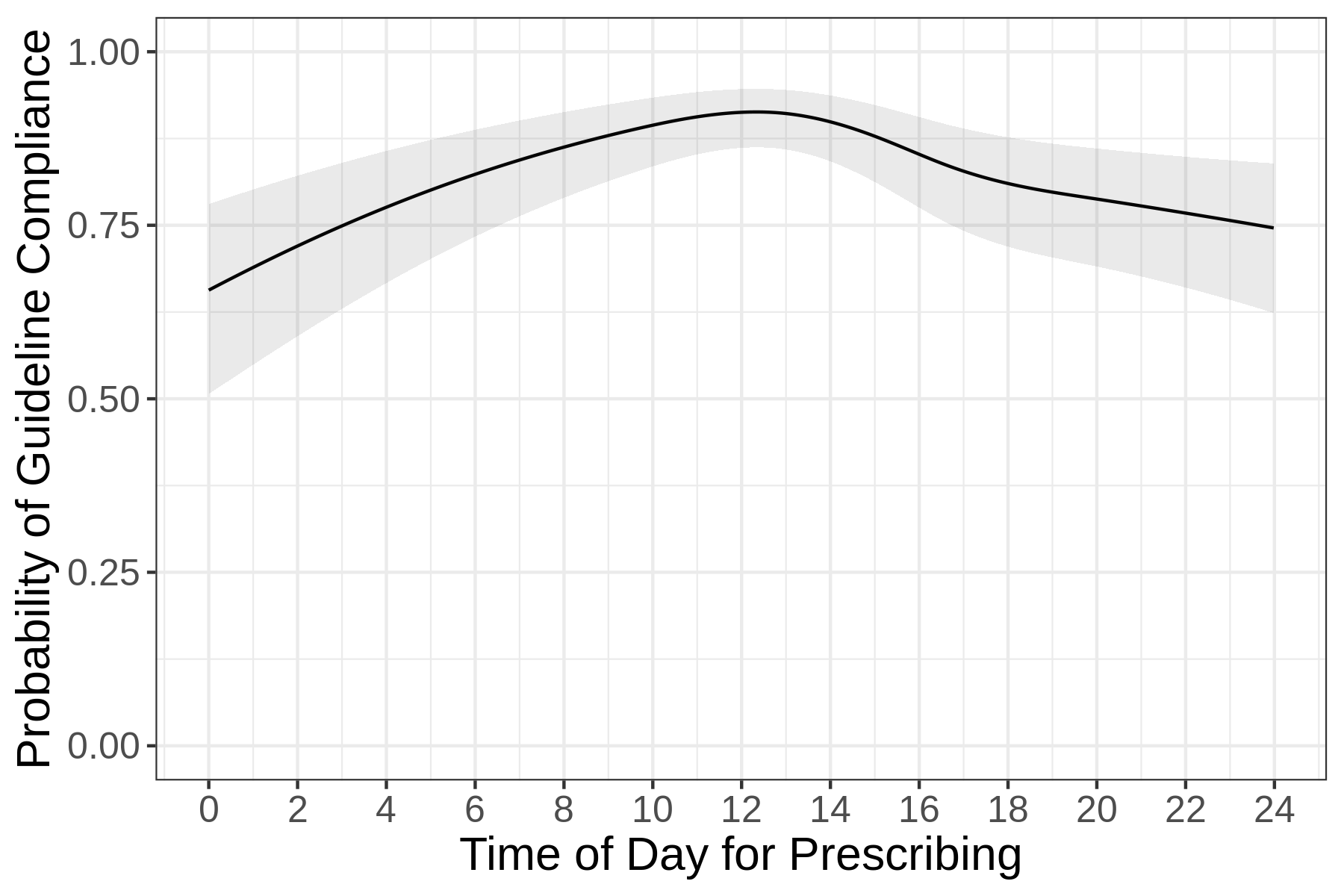

**Figure S2**. **Associations between time of day of the prescription and guideline compliance of the loading dose.** Other predictors were held constant at their mean (for continuous variables) or reference levels (for categorical variables).

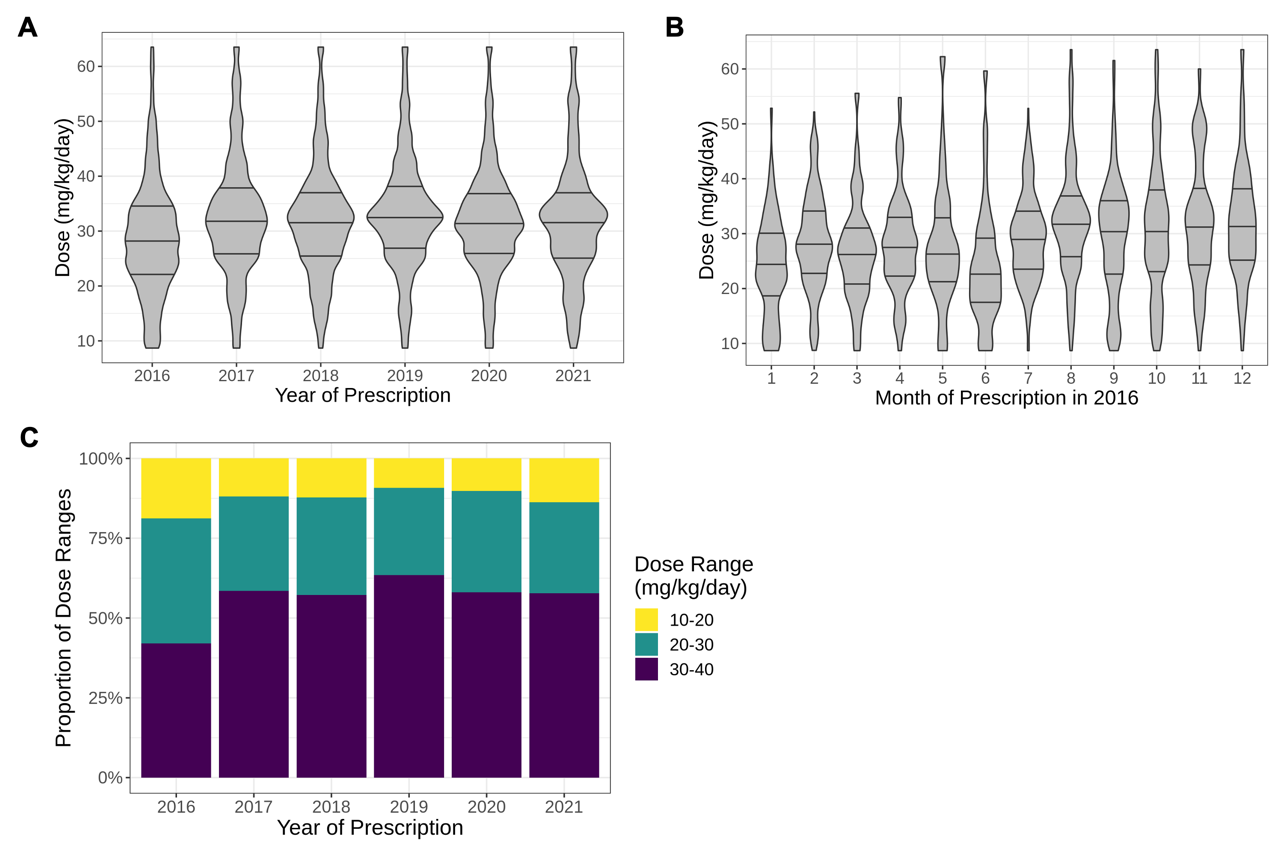

**Figure S3**. **Subsequent vancomycin maintenance doses.** Panel A shows doses by years between 2016 and 2021, and panel B by month in 2016. Panel C shows the proportion of doses in three dose ranges (10–20mg/kg/day., 20–30mg/kg/day, 30–40mg/kg/day) over 2016-2021.

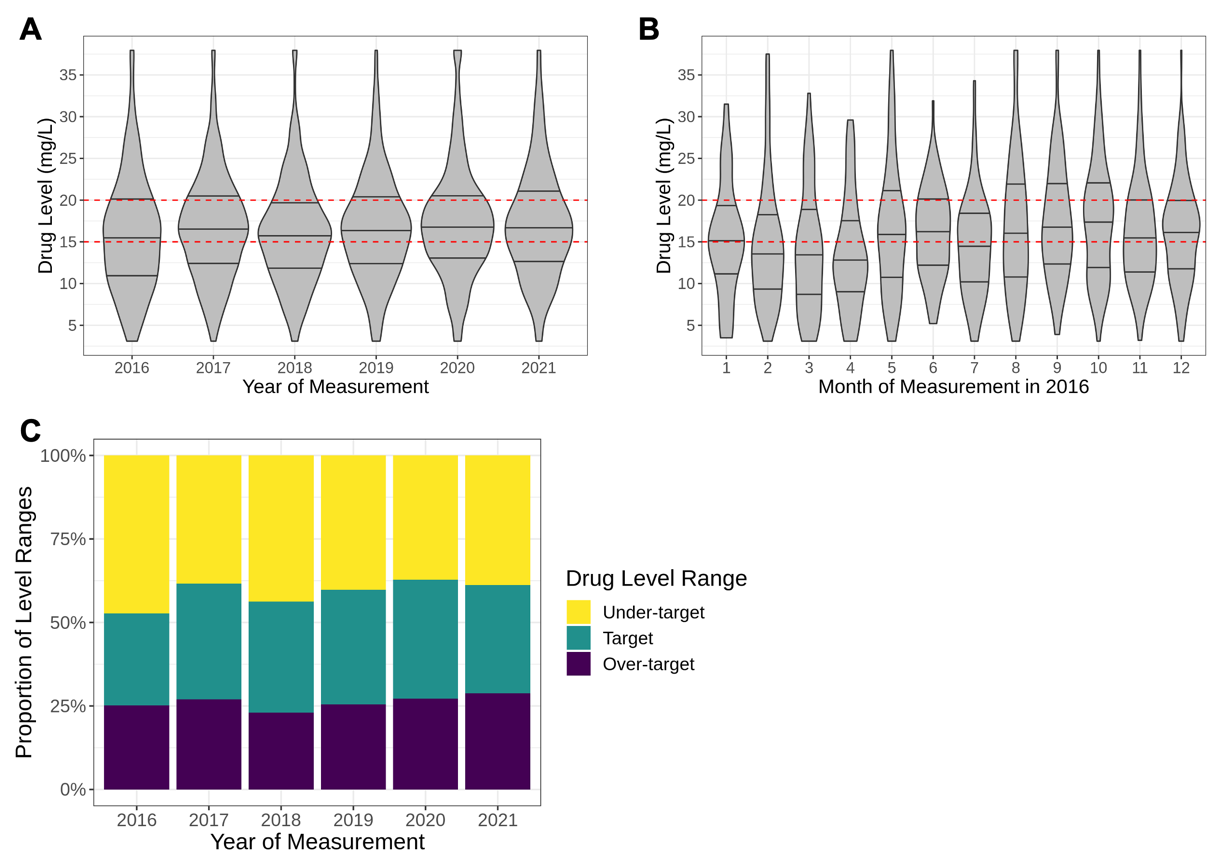

**Figure S4. Subsequent vancomycin drug levels.** Panel A shows drug levels by years between 2016 and 2021, panel B by months in 2016. Panel C shows the proportion of levels in different drug level ranges (under-target, target, over-target) over 2016-2021. The dashed red lines in panels A and B indicate the target drug range (15–20 mg/L).

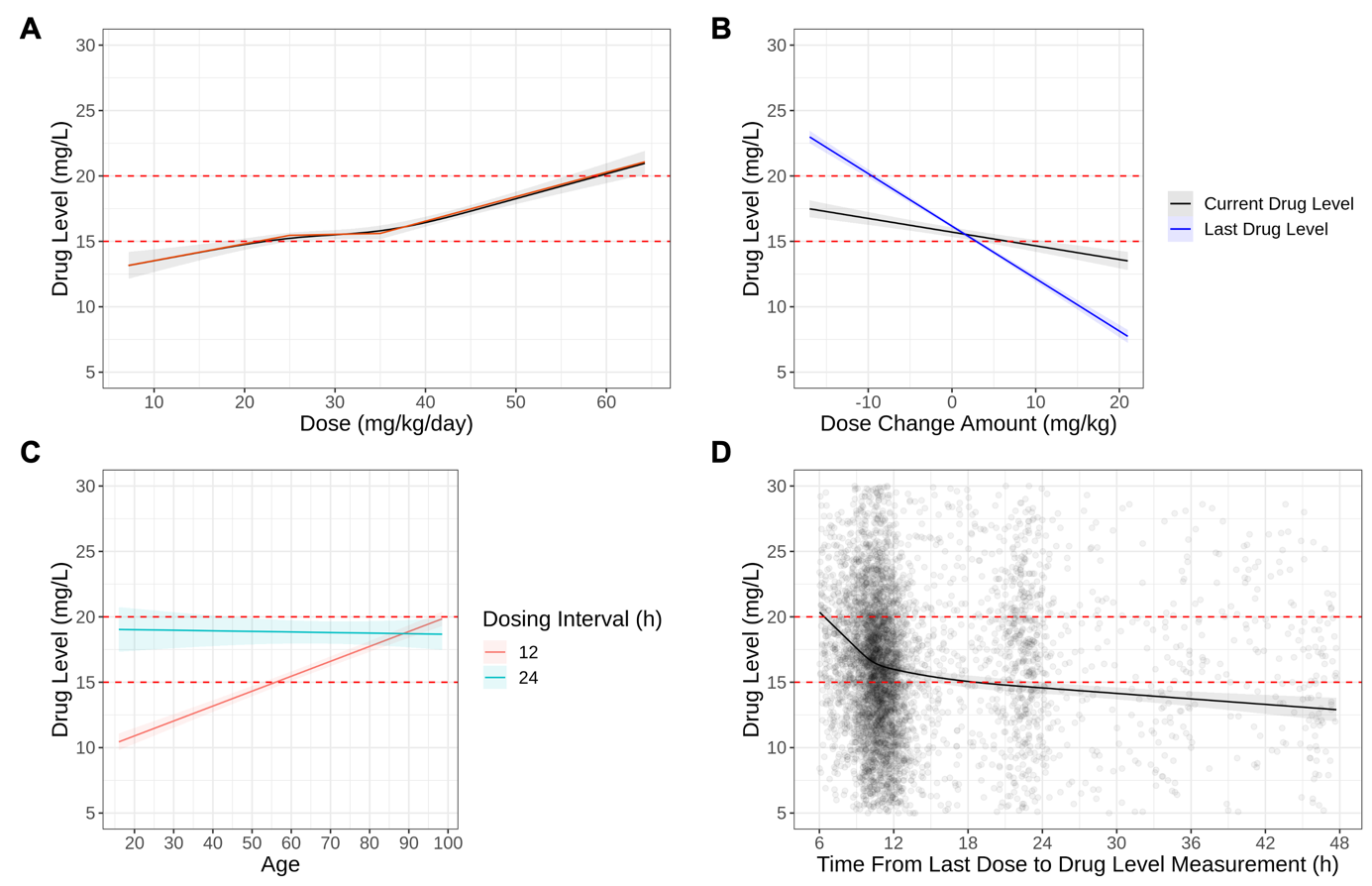

**Figure S5. The associations between subsequent drug levels and post-initial maintenance doses (panel A), dose change (panel B), age (panel C), time from the last dose to drug level measurement (panel D).** (A) Non-linear associations between drug levels and subsequent doses (black line) are approximated with piecewise linear splines (red line). (B) The blue line represents the drug level measured before dose adjustment. (D) The data points represent the actual time of blood sampling versus drug levels. The red dashed line represents the target trough level range recommended in the dosing guideline (15-20mg/L). Other predictors were held constant at their mean (for continuous variables) or reference levels (for categorical variables).

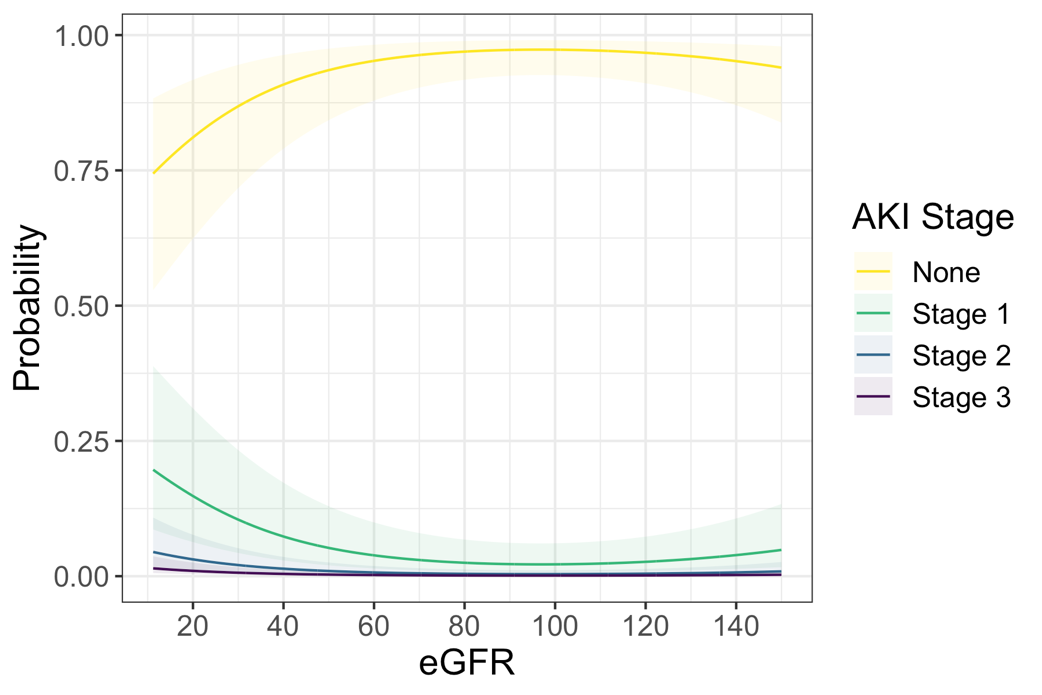

**Figure S6.** **The associations between the probability of different stages of AKI and pre-treatment eGFR.** Other predictors were held constant at their mean (for continuous variables) or reference levels (for categorical variables).
